# Safety of the BNT162b2 mRNA COVID-19 Vaccine in Children below 5 Years (CoVacU5) – an Investigator-Initiated Retrospective Cohort Study

**DOI:** 10.1101/2022.05.17.22275005

**Authors:** Nicole Töpfner, Wolfgang C G von Meißner, Christoph Strumann, Denisa Drinka, David Stuppe, Maximilian Jorczyk, Jeanne Moor, Johannes Püschel, Melanie Liss, Emilie von Poblotzki, Reinhard Berner, Matthias B. Moor, Cho-Ming Chao

## Abstract

**Background:** The safety of SARS-CoV-2 vaccines is unknown in children aged <5 years. Here, we retrospectively evaluated the safety of BNT162b2 vaccine used off-label in children of this age group in Germany.

**Methods:** An investigator-initiated retrospective cohort study (CoVacU5) included parents or caregivers having children aged <5 years registered for SARS-CoV-2 vaccination in outpatient care facilities in Germany. Reported short-term safety data of 1-3 doses of 3-10µg BNT162b2 in children aged 0 to <60 months are presented. Co-primary outcomes were the frequencies of 11 categories of symptoms post-vaccination with bivariate analyses and regression models adjusting for age, sex, weight and height. On-label non-SARS-CoV-2 vaccines served as controls in an active-comparator design.

**Results:** The study included 7806 of 19,000 registered children representing a 41% response rate. 338 children received the first dose of BNT162b2 at age 0-<12 months, n=1272 at age 12-24 months and n=5629 at age ≥24 to <60 months. A 10µg dosage was more frequently associated with injection-site symptoms compared to lower dosages. The probability of any symptoms (OR: 1.62 [95% confidence interval (CI): 1.36-1.94]), injection-site, musculoskeletal, dermatological or otolaryngological symptom categories were modestly elevated after BNT12b2 compared to non-SARS-CoV-2 vaccines, whereas the probabilities of general symptoms (OR: 0.74 [95% CI: 0.64-0.85]) and fever (OR: 0.43 [95% CI: 0.35-0.51]) were lower after BNT162b2. Symptoms requiring hospitalization (n=10) were reported only at BNT162b2 dosages higher than 3µg.

**Conclusions:** The symptoms reported after BNT162b2 administration were overall comparable to on-label non-SARS-CoV-2 vaccines in this cohort of children aged <5 years. (German Clinical Trials Register ID: DRKS00028759).

## Introduction

The safety and efficacy of the SARS-CoV-2 mRNA vaccine BNT162b2 (Comirnaty^®^) has been studied in over 2500 children and adolescents aged ≥5 years (1–3). However, in children below 5 years the use of SARS-CoV-2 vaccines is currently not approved by the European Medicines Agency, while studies evaluating SARS-CoV-2 vaccines in this age group are ongoing (4,5). Public news media have reported since mid-2021 that German initiatives by laypeople or medical care providers have organized off-label SARS-CoV-2 vaccinations (6–9), including children who or whose family members have comorbidities or medications that may increase their risk for severe COVID-19. Off-label administration of SARS-CoV-2 vaccines to children aged below 5 years is permitted according to German law after obtaining informed consent, but remains at parents’, legal guardians’ or healthcare providers’ risk or liability (10,11). Parents or legal guardians may seek opportunities to vaccinate children against SARS-CoV-2 in an attempt to obtain according to their individual estimation the best attainable protection against COVID-19 for their child and/or potentially diminish the risk of post-acute syndromes or virus transmission. On February 17^th^ 2022, the German government’s COVID-19 advisory council has stated that children’s welfare in times of pandemic requires prioritization, including systematic efforts to study potential SARS-CoV-2 vaccinations side effects (12,13). At present, no safety data are available for BNT162b2 in children aged <5 years.

## Methods

### Study design

The CoVacU5 study was designed as a retrospective cohort study to evaluate the safety of BNT162b2 mRNA vaccine in children vaccinated before completing the 5^th^ year of age. Inclusion criteria were having registered in a database for at least one dose of BNT162b2 that was administered before reaching the age of 5 years, and informed consent of a parent/legal representative to participate in the survey of the current study. Eligible participants were identified from electronic registration databases by 21 outpatient care facilities and two nation-wide layperson-initiated SARS-CoV-2 vaccination programs in Germany. Exclusion criteria were duplicate responses with overlapping age, sex, weight or height without specification that they were twins or triplets, and respondents lacking an authentication code as proof of invitation.

The present study protocol was approved by the Ethics Committee of the University of Rostock, Germany (ID: A 2022-0065). The study was prospectively registered in the German Clinical Trials Register (ID: DRKS00028759) and was conducted with adherence to the Declaration of Helsinki. The authors vouch for the fidelity to the approved study protocol. Parents or legal representatives of all studied children gave informed consent for the participation in the retrospective evaluation in the present survey. Of note, the vaccines were neither administered as part of the study nor was the study planned or initiated before vaccination. Parents or legal representatives of the studied children had independently given individual informed consent to the vaccinations and had been informed by the attending physicians about potential side effects and liability of this off-label medicine use under the German law. The attending physicians were themselves required by German Infection Protection Act and by their professional status to report all unexpected or severe side effects experienced by the children to the German Federal Institute for Drugs and Medical Devices independently from the present study.

Within the study period between April 14^th^ and May 9^th^ 2022, the parents’ or caregivers were contacted twice via e-mail addresses registered in the databases and were invited to participate in a web-based survey. All invitees received an 8-digit authentication code to ensure participation of only those who had effectively registered at least one child for vaccination. On May 9^th^, 2022 the study database was closed and data were extracted for analysis in a fully anonymous manner.

### Study procedure

All study data were collected and managed using REDCap (14) electronic data capture tools hosted at the University Medical Center Dresden, Germany. The survey was designed to be completed within approximately 5 minutes in case no symptoms were to be reported, and it was pre-tested in 20 pilot test runs. An introductory page revealed the study purpose and obtained informed consent for voluntary participation and allowing data use for research purposes. Subsequently, demographic information, comorbidities and medications of the children were collected, and precise symptoms including their timing post-vaccination were determined by close-ended questions and free-text choices. The symptom categories were vaccination-site symptoms, general reactions, fever, skin changes, or any symptoms of the musculoskeletal, cardiovascular, pulmonary or nervous system, gastrointestinal tract, or, as well as any symptoms of ear/neck/throat, psychological symptoms or susceptibility for infections. In addition, the occurrence of missing days from childcare institution or school was recorded. An item based on a 10-point Likert scale was used to determine if the reported post-vaccination symptoms in the children were considered to be medically threatening. For comparison, information was collected regarding post-vaccination symptom categories of any other non-SARS-CoV-2 vaccination administered in the same children since January 15^th^, 2022.

### Outcomes

Primary outcome of the present study was the probability of self-reported categorized symptoms after BNT162b2 in comparison to non-SARS-CoV-2 vaccines with particular respect to SAE, and the frequencies of categorized symptoms occurring after BNT162b2 administration with stratification by age group and dosage, and the general susceptibility to infections and institutional missing days.

Secondary outcomes were the total number, duration and consequences of symptoms occurring after vaccination with BNT162b2.

### Statistical analyses

All eligible participants were enrolled without sample size calculations. Statistical analyses were performed using MATLAB R2020a and STATA version 15. Chi-Square or Fisher’s exact tests were used to compare categorical variables over pre-defined strata of age groups and BNT162b2 dosage. T-test or Wilcoxon rank sum tests were used for comparisons of continuous variables as appropriate. Multivariable logistic models were performed with any symptom or the 11 individual categories of post-vaccination symptoms as dependent variables, with BNT162b2 versus non-SARS-CoV-2 vaccine as predictor of interest. These models were adjusted for age, sex, weight and height. Duration and impact of the symptoms were assessed in additional logistic models. In addition, multivariate negative binomial regression was used for the number of symptoms as dependent variable. For bivariate analyses and regression models, p values and 95% confidence intervals were adjusted for multiple testing of 11 symptom categories using the Bonferroni method. An adjusted p<0.05 (signifying an unadjusted p<0.004) was considered significant. A robustness check of the data was performed by multiple imputation. Complete variables serving as explanatory variables were included in the imputation models. Additional, aggregated geolocation dummy variables based on the postal code were used as regression variables in the imputation models if they obtained complete data. As a sensitivity analysis, participants who did not provide a verifiable (15) lot number of BNT162b2 vaccinations were excluded.

## Results

### Study population

In the present study, the self-reported safety of BNT162b2 mRNA vaccine against SARS-CoV-2 administered off-label to children aged 0 to <5 years in outpatient care facilities in Germany was analyzed.

Figure 1 shows the flowchart of participant recruitment. Participants were invited via an estimated 19,000 e-mail addresses available by vaccine registration databases of 21 outpatient care facilities and the initiatives “U12Schutz” and “Bildung Aber Sicher” in Germany. This led to a total of 11,542 responses. After exclusion of 3209 respondents providing empty datasets, 173 respondents with children aged ≥5 years at first BNT162b2 vaccine administration, and 117 respondents having entered no valid authentication code, 8043 responses remained. From these, 119 potential duplicate entries were removed based on identical demographic data, additionally 49 respondents with BNT162b2 vaccinations reported to have occurred prior to May 2021, and 69 respondents reporting vaccinations with mRNA-1273 (Moderna). This yielded a final population of 7806 participants included in the present analysis, representing a response rate of 41%. We included 338 children aged <12 months, 1272 children aged 12 to <24 months and 5629 children aged 24 to <60 months at the age of their first BNT162b2 vaccination, some of whom were aged above 5 years at their second or third BNT162b2 vaccination. The present analysis included at least 1 vaccine dose administered in 7806, at least 2 doses in 7102 and 3 doses in 846 children. For 7750 of the 15,754 (49.2%) vaccinations with BNT162b2, manufacturer-verified lot numbers were reported. Population characteristics, dosage of administered vaccines, and interval between vaccines are shown in Table 1. A total of 4.9% reported regular medication use for underlying diseases and 8.8% reported existing comorbidities (Table 1). Among the most common comorbidities were pulmonary (2.4%) or cardiovascular diseases (2.1%) and chromosomal aberrations (1.2%) (Supplementary Table 1).

**Figure 1.**
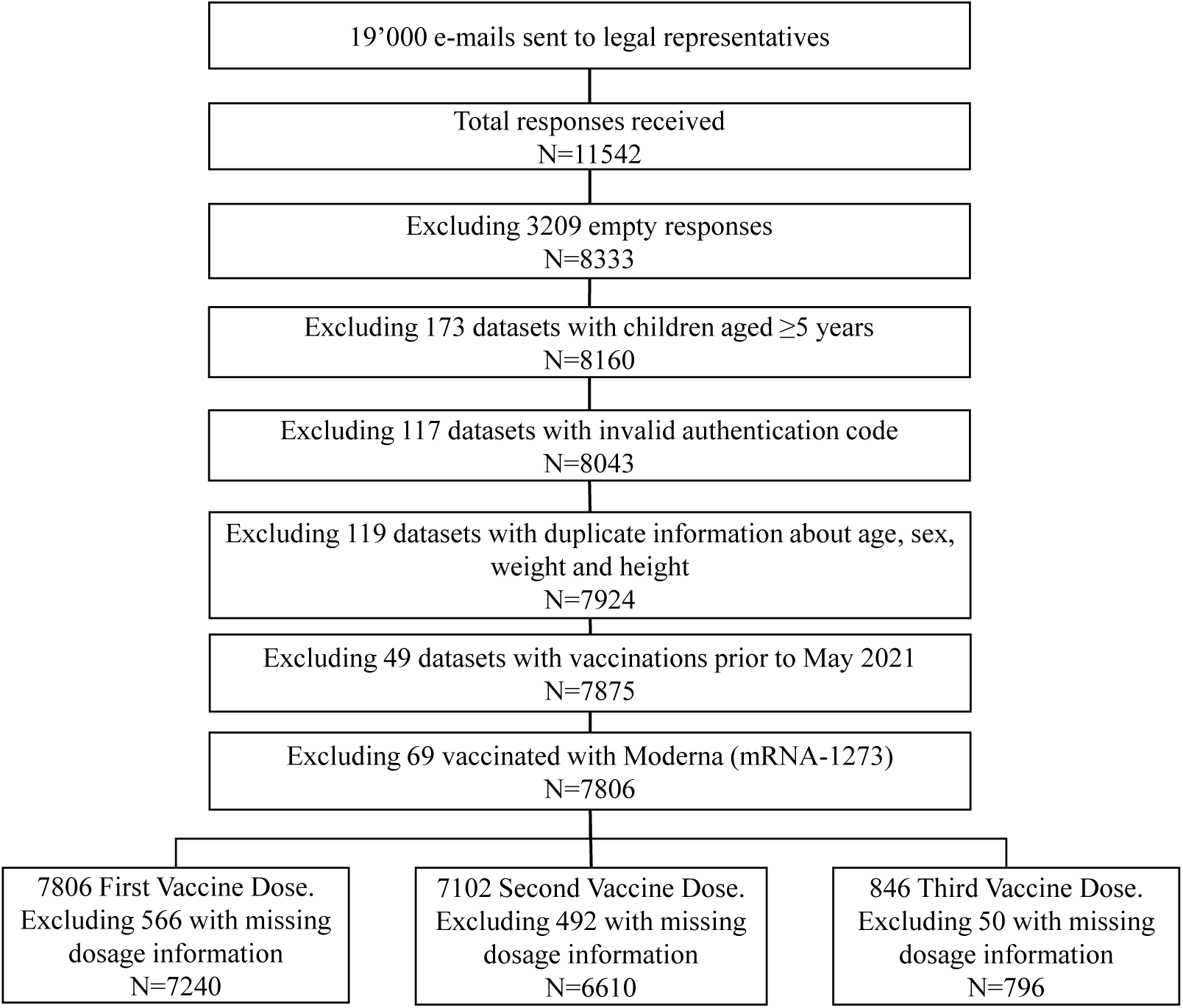
Overview of eligible and enrolled study participants. The diagram displays the number of participants contacted via e-mail, all excluded responses and the final number of BNT162b2 vaccine administrations analyzed.

**Table 1:** Cohort Characteristics, n/N(%)

|  | All | First<br>vaccination | Second<br>vaccination | Third<br>vaccination |
| --- | --- | --- | --- | --- |
| Female | 3824/7805 (49) | 3824/7805 (49) | 3482/7101 (49) | 419/846 (49.5) |
| Male | 3977/7805 (51) | 3977/7805 (51) | 3616/7101 (50.9) | 427/846 (50.5) |
| Diverse | 4/7805 (0.1) | 4/7805 (0.1) | 3/7101 (0) | 0/846 (0) |
| Age (years) (median,<br>IQR) | 3 (2) | 3 (2) | 3 (2) | 3 (1.5) |
| Height (cm) (median,<br>IQR) | 98 (17) | 98 (17) | 98 (17) | 99 (13) |
| Weight (kg) (median,<br>IQR) | 14.5 (5) | 14.5 (5) | 14.5 (5) | 15 (4.5) |
| BNT162b2: 3µg | 2995/14646 (20.4) | 1772/7240 (24.5) | 1171/6610 (17.7) | 52/796 (6.5) |
| BNT162b2: 5µg | 6075/14646 (41.5) | 3120/7240 (43.1) | 2740/6610 (41.5) | 215/796 (27) |
| BNT162b2: 10µg | 5576/14646 (38.1) | 2348/7240 (32.4) | 2699/6610 (40.8) | 529/796 (66.5) |
| Interval between the<br>last vaccination (mean<br>days) |  |  | 25.8 | 79.4 |
| Comorbidities (yes) | 684/7806 (8.8) | 684/7806 (8.8) | 613/7102 (8.6) | 89/846 (10.5) |
| Long-term<br>Medication (yes) | 381/7784 (4.9) | 381/7784 (4.9) | 337/7099 (4.7) | 46/846 (5.4) |

IQR, interquartile range.

### Frequency of post-vaccination symptoms after BNT162b2 administration

A descriptive analysis of symptoms reported after vaccination was performed. Frequencies of all reported symptom categories occurring after first or second BNT162b2 vaccination are shown in Figure 2 and Supplementary Table 2-3, and for the third BNT162b2 vaccination in Supplementary Table 4, with stratification for age groups and BNT162b2 vaccine dosages. While most categories were comparable across the strata, local injection-site symptoms were more frequently reported for the age group ≥24-<60 months than for younger children. After the third vaccination, too few data were available to allow a conclusive comparison of symptom frequency e.g. between the dosages, but a non-significant trend towards dose-dependent local reactions was present (Supplementary Table 4). No significant age-dependent effects of reported symptoms, and no drug dose-dependent effects with the exception of dose-dependent vaccination-site symptoms were found in 24 to <60 months old children. The vast majority of all BNT162b2 post-vaccinations symptoms was judged of no or minimal subjective severity and did not lead to any missing days in school or childcare (Supplementary Table 5). We retrospectively defined post-vaccination mortality (n=0/7806, 0%) and symptoms requiring inpatient treatment (n=10/7806, 0.1%) as serious adverse events (SAE). We additionally considered symptoms as of concern when they lasted over 3 months (n=2/7806 respondents, 0%) and of unknown significance when they were currently ongoing (n=40/7806, 0.5%). Symptoms of the 10 children with SAE lasted 12.2±18.6 days (mean +-standard deviation) [maximum: 60 days] and were observed after BNT162b2 administration at 5µg, 10µg or unknown/other dosage, but none after the 3µg dosage (Table 2) and none after the third vaccination with BNT162b2. A full analysis of post-vaccination symptoms including time of onset, duration and perceived severity is shown for all individual symptom categories in Supplementary Tables 6-15.

**Figure 2.**
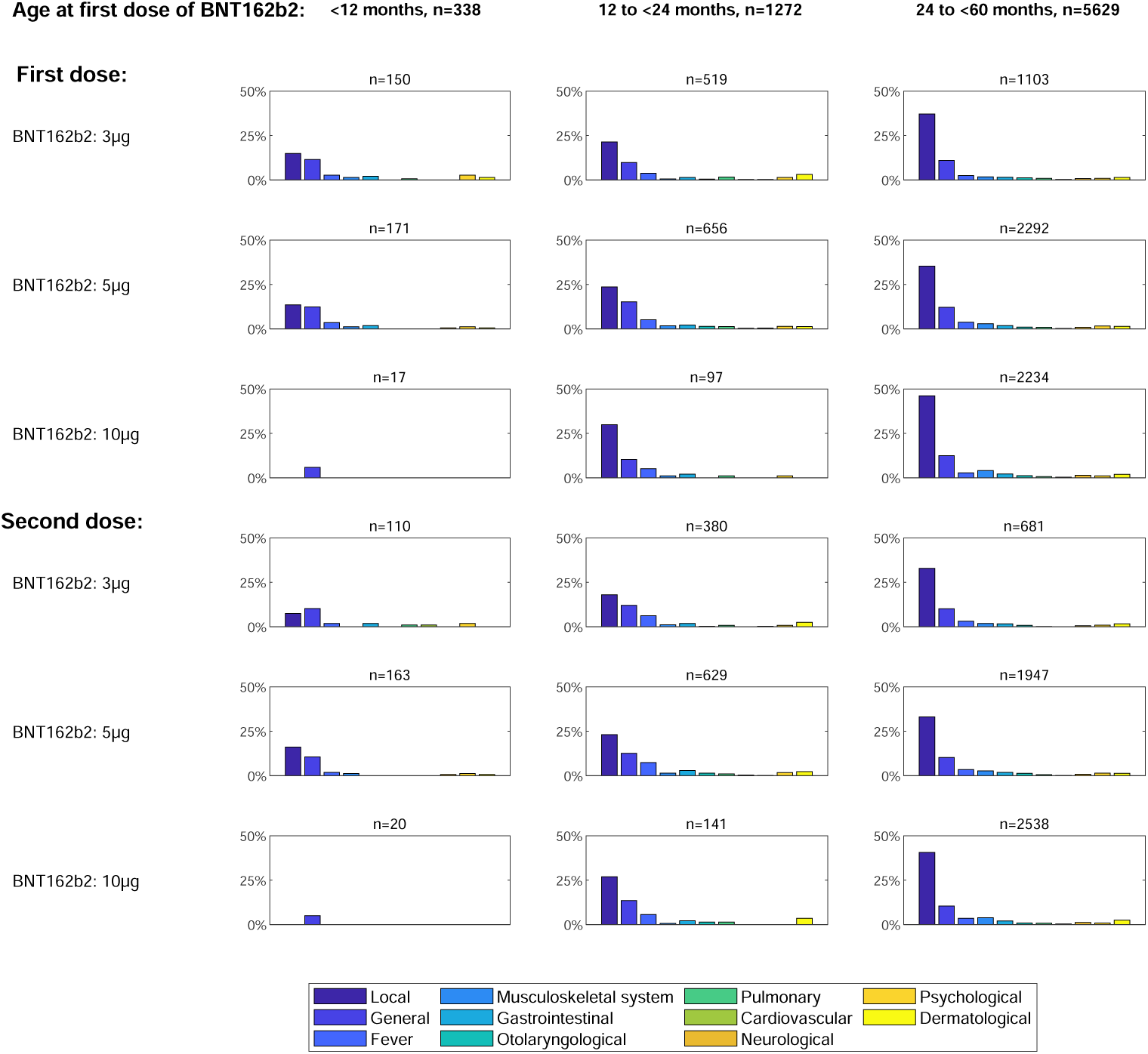
Symptoms reported after BNT162b2 vaccine administration according to age group and dosage. Frequencies of retrospectively reported symptom categories occurring after the first vaccination (upper panel) or the second vaccination (lower panel) with BNT162b2.

**Table 2:** Reported serious adverse events after BNT162b2 administration in 7806 children under the age of 5 years

|  | Overall | First<br>BNT162b2<br>vaccination | Second<br>BNT162b2<br>vaccination |
| --- | --- | --- | --- |
| N(%) | 10(100) | 10(100) | 9(90) |
| Female | 7(70) |  |  |
| Male | 3(30) |  |  |
| Age (years), median (IQR) | 4(1) |  |  |
| Comorbidities (Yes) | 6(60) |  |  |
| Long-term Medication | 2(20) |  |  |
| Inpatient treatment | 10(100) | 7(70) | 3(30) |
| *Dosage* |  |  |  |
| 3µg |  | 0(0) | 0(0) |
| 5µg |  | 3(30) | 4(40) |
| 10µg |  | 5(50) | 5(50) |
| unknown |  | 2(20) | 1(10) |
| *Symptoms* |  |  |  |
| Duration (days), mean±std, max | 12.2±18.6,60 | 4.3±5.4,14 | 9.3±20.8,60 |
| Duration > 90 days | 0(0) | 1(10) | 0(0) |
| Duration ongoing | 1(10) | 0(0) | 0(0) |
| Local | 5(50) | 5(50) | 3(33.3) |
| General | 5(50) | 3(30) | 3(33.3) |
| Fever | 2(20) | 2(20) | 0(0) |
| Musculoskeletal system | 4(40) | 2(20) | 3(33.3) |
| Gastrointestinal | 1(10) | 0(0) | 1(11.1) |
| Otolaryngological | 3(33.3) | 2(22.2) | 1(12.5) |
| Pulmonary | 4(40) | 4(40) | 0(0) |
| Cardiovascular | 4(40) | 2(20) | 3(33.3) |
| Neurological | 3(30) | 1(10) | 2(22.2) |
| Psychological | 1(10) | 0(0) | 1(11.1) |
| Dermatological | 2(20) | 1(10) | 1(11.1) |

IQR, interquartile; std, standard deviation.

### Active-comparator analysis of BNT162b2 versus non-BNT162b2 vaccines administered between January 15^th^ and May 9^th^, 2022

Subsequently, a comparison of post-vaccination symptoms of BNT162b2 in off-label use with those occurring after on-label vaccinations against pathogens other than SARS-CoV-2 was performed as displayed in Supplementary Table 16 within the same group of participants. To this end, the symptoms of all BNT162b2 or non-SARS-CoV-2 vaccinations administered in the time period between January 15^th^, 2022 and May 9^th^, 2022 were assessed to minimize potential differences in recall bias between vaccine groups. Any symptoms were observed after 50.8% of BNT162b2 and 37.8% of non-SARS-CoV-2 vaccinations. In logistic regression models adjusted for age, sex, weight and height, BNT162b2 vaccination was associated with a higher probability of any reported post-vaccination symptom (OR: 1.62 [95% CI: 1.36;1.94], p<0.001) in comparison to non-BNT162b2 vaccination (Table 3). Similarly, the probabilities of musculoskeletal, otolaryngological or dermatological symptoms were higher after BNT162b2 compared to non-SARS-CoV-2 vaccination. However, the probabilities of general symptoms or fever were lower after BNT162b2 vaccinations compared to non-SARS-CoV-2 vaccination (OR: 0.77 [95% CI: 0.63;0.95], p<0.001 and OR: 0.42 [0.32;0.55], p<0.001 respectively). The probabilities of all other symptom categories were comparable between BNT162b2 and non-SARS-CoV-2 vaccinations (Table 2). A symptom duration over 3 months was reported after 1/4570 (0%) of BNT162b2 and none of non-SARS-CoV-2 vaccinations, symptoms currently ongoing after 24/4570 (0.5%) of BNT162b2 and 1/1491 (0.1%) of non-SARS-CoV-2 vaccinations, and symptoms requiring inpatient treatment were reported after 5/4570 (0.1%) of BNT162b2 and none of non-SARS-CoV-2 vaccinations.

**Table 3:**
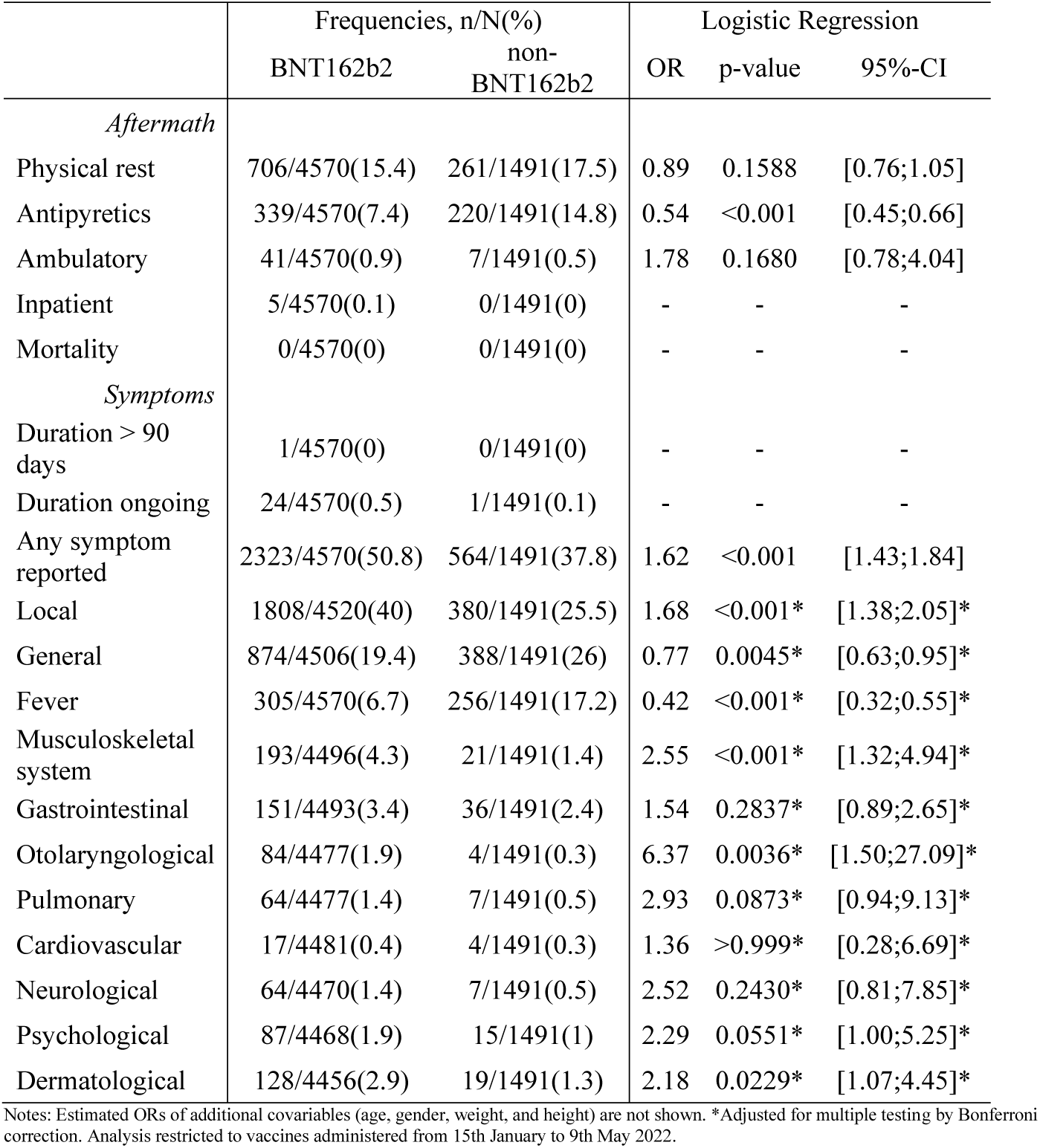
Active-comparator analysis of symptoms occurring after BNT162b2 and non-BNT162b2 vaccinations

|  | Frequencies, n/N(%) |  | Logistic Regression |  |  |
| --- | --- | --- | --- | --- | --- |
|  | BNT162b2 | non-BNT162b2 | OR | p-value | 95%-CI |
| *Aftermath* |  |  |  |  |  |
| Physical rest | 706/4570(15.4) | 261/1491(17.5) | 0.89 | 0.1588 | [0.76;1.05] |
| Antipyretics | 339/4570(7.4) | 220/1491(14.8) | 0.54 | <0.001 | [0.45;0.66] |
| Ambulatory | 41/4570(0.9) | 7/1491(0.5) | 1.78 | 0.1680 | [0.78;4.04] |
| Inpatient | 5/4570(0.1) | 0/1491(0) | - | - | - |
| Mortality | 0/4570(0) | 0/1491(0) | - | - | - |
| *Symptoms* |  |  |  |  |  |
| Duration > 90 days | 1/4570(0) | 0/1491(0) | - | - | - |
| Duration ongoing | 24/4570(0.5) | 1/1491(0.1) | - | - | - |
| Any symptom reported | 2323/4570(50.8) | 564/1491(37.8) | 1.62 | <0.001 | [1.43;1.84] |
| Local | 1808/4520(40) | 380/1491(25.5) | 1.68 | <0.001* | [1.38;2.05]* |
| General | 874/4506(19.4) | 388/1491(26) | 0.77 | 0.0045* | [0.63;0.95]* |
| Fever | 305/4570(6.7) | 256/1491(17.2) | 0.42 | <0.001* | [0.32;0.55]* |
| Musculoskeletal system | 193/4496(4.3) | 21/1491(1.4) | 2.55 | <0.001* | [1.32;4.94]* |
| Gastrointestinal | 151/4493(3.4) | 36/1491(2.4) | 1.54 | 0.2837* | [0.89;2.65]* |
| Otolaryngological | 84/4477(1.9) | 4/1491(0.3) | 6.37 | 0.0036* | [1.50;27.09]* |
| Pulmonary | 64/4477(1.4) | 7/1491(0.5) | 2.93 | 0.0873* | [0.94;9.13]* |
| Cardiovascular | 17/4481(0.4) | 4/1491(0.3) | 1.36 | >0.999* | [0.28;6.69]* |
| Neurological | 64/4470(1.4) | 7/1491(0.5) | 2.52 | 0.2430* | [0.81;7.85]* |
| Psychological | 87/4468(1.9) | 15/1491(1) | 2.29 | 0.0551* | [1.00;5.25]* |
| Dermatological | 128/4456(2.9) | 19/1491(1.3) | 2.18 | 0.0229* | [1.07;4.45]* |

Notes: Estimated ORs of additional covariables (age, gender, weight, and height) are not shown. \*Adjusted for multiple testing by Bonferroni correction. Analysis restricted to vaccines administered from 15th January to 9th May 2022.

### Sensitivity analyses

The present dataset contained a number of less stringently verifiable participants that revealed no lot number of the BNT162b2 vaccine. Thus, we performed a sensitivity analysis and reanalyzed the data presented in Supplementary Tables 2-4 and Table 3 excluding all those with missing data in lot numbers. These analyses yielded no differences in results (data not shown). Similarly, multiple imputation of missing data (Supplementary Table 17) followed by re-analysis of the data presented in Supplementary Tables 2-4 and Table 3 did not affect the results (data not shown).

## Discussion

The CoVacU5 study is an industry-independent retrospective cohort study designed to assess the self-reported short-term safety of BNT162b2 vaccines in young children in a setting where parents or caregivers had decided beforehand and independently from this study to seek off-label vaccination of their entrusted children. This study included the BNT162b2 vaccination experience of 7806 children. Due to the large study population and the different dosages investigated, CoVacU5 expands the available knowledge of mRNA vaccination in children aged <5 years in addition to the results of the ongoing industry-sponsored BNT162b2 phase I to III studies conducted in this age group (5).

This study revealed a similar overall safety when compared with existing non-SARS-CoV-2 vaccines administered in the same study population, with minor differences in the probabilities of injection-site symptoms, fever, musculoskeletal, or otolaryngological symptoms in comparison to non-SARS-CoV-2 vaccinations. An increased frequency of injection-site symptoms was detected in ≥24 to <60 months old children administered 10µg of BNT162b2, which should be taken into consideration in future dosage-finding strategies in this age group and be carefully weighed against potential improvements in immunogenicity at higher dosage. In 40 individuals symptoms were currently ongoing and thus of unknown significance. In two individuals, symptoms lasted longer than 90 days, and in 10 children hospitalizations were reported which were considered SAE. Although the circumstances of these SAE are not precisely documented, no SAE were reported for children administered the low dosage of 3µg that is currently evaluated in the ongoing BNT162b2 licensure study (5). In addition, no mortality was reported.

A relevant support for the reliability of our study result is that all physicians who administer vaccines in Germany are obliged by Federal Law to report all severe or unexpected vaccine-related side effects to the Federal Institute for Vaccines and Biomedicines (Paul-Ehrlich Institute). In addition to the here presented data, the Paul-Ehrlich Institute had not received any reports of unexpected or severe adverse reactions of BNT162b2 in children aged <5 years as of March 31^st^, 2022 (16).

The present study contains some limitations. First, the study relies on retrospective self-reported data by proxy, which i) may not directly reflect what a child had experienced and ii) underlies a risk of recall bias i.e. not recalling some not life-threatening symptoms several months later. However, the extensive public debate about SARS-CoV-2 vaccinations including their potential side effects, the off-label use investigated, and the study results showing dosage-dependent associations between vaccination and local reactions in ≥24 to <60 months old children suggest that symptoms were adequately reported. The retrospective design did not allow for a structured real-time documentation of symptoms nor an estimation of causality and severity to infer the frequency of vaccine-related serious adverse reactions, but such data should be expected from the ongoing prospective manufacturer-sponsored BNT162b2 studies (5). As vaccinations themselves were not directly part of the study, the study relied on vaccination information as reported by the respondents. However, a potential risk of manipulation by individual respondents was minimized by dispensing study participation codes only via vaccinating centers/initiatives, and the availability of BNT162b2 lot numbers. Moreover, potential duplicate entries were identified by overlap in demographic data and were removed.

The reported post-vaccination symptoms could potentially be unrelated to the vaccine. To mitigate this problem, a non-BNT162b2 vaccination group as an active comparator was included that yielded an overall similar probability of post-vaccination symptoms for most symptom categories within the last three months. In the timeframe of this analysis, vaccinations from January 15^th^ to May 9^th^ 2022 concurred with the peak of Omicron BA.1 and BA.2 waves, leading to potentially some symptoms being caused by undiagnosed SARS-CoV-2 but not any of the vaccines. This affects both analyzed groups similarly and does unlikely bias the observed findings.

Finally, a particular focus has to be dedicated to the fact that the vaccines administered in the present population were used off-label. Although off-label use is common in pediatric care, constituting up to 25% of all medications administered to children (17), administration of a new vaccine in off-label use constitutes an even more particular situation. There was extensive public discussion on SARS-CoV-2 vaccination of children in Germany even in age groups where the vaccine had been licensed. Off-label vaccination of the most vulnerable population of very young children is an even more challenging and provoking issue. However, since it was publicly known that a large group of young children in Germany is being vaccinated by the active and autonomous decision of their parents or legal guardians, it appeared to be responsible and important from a scientific as well as from an ethical point of view to collect all available information, to analyze these data in a scientifically sound matter, and to make this information available to the medical community as well as to the public. Of course, this was not a prospective, randomized and controlled study design and the quality of the data is limited by nature. Nevertheless, the scientific use and thorough evaluation of existing, retrospectively self-reported safety data for SARS-CoV-2 vaccines in off-label use appeared mandatory, and may – respecting all given limitations – expand the scientific knowledge based on real-world medical practice.

## Conclusion

The study data provide evidence for a self-reported safety profile of BNT162b2 vaccine that is comparable to non-SARS-CoV-2 vaccines in this large cohort of children aged <5 years until today. These data can be helpful in safety considerations for individual decision-making and may add to data expected from prospective licensure studies for expert recommendations about BNT162b2 vaccinations in this age group, even after the completion of the ongoing phase I/II/III study of BNT162b2 (5).

## Data Availability

The raw data are available from the corresponding author on reasonable request, provided that a positive evaluation by the pertinent ethics committee is available and German data protection laws are followed.

## Author contributions

CMC conceived the study. NT, WCGM, JM, MBM and CMC designed the study. NT, DD, DS, MJ and CMC designed the questionnaire with inputs from WCGM, JM and MBM. CS and MBM designed the statistical analysis plan with inputs from NT and CMC. WCGM, DS, JP, ML, EP and CMC recruited participants. CS performed analyses. NT, WCGM, CS, DS, RB, MBM and CMC interpreted the data. MBM wrote the manuscript with inputs from all other authors. NT, WCGM, JM, RB, MBM and CMC edited the manuscript. All authors approved the final version of the submitted manuscript.

## Acknowledgements

The authors are thankful to all participating vaccination centers and respondents. We acknowledge the non-profit assistance for participant recruitment provided by the initiatives “Bildung Aber Sicher”, “U12Schutz” and by the individuals Anke Böhnke, Armin Philipp, Georg Hillebrand, Lisa Degener, and Lisa Schneider.

## Registration

The present work was registered at the German Clinical Trials Register ID: DRKS00028759.

