## Supplementary Table for "Safety of the BNT162b2 mRNA COVID-19 Vaccine in Children below 5 Years (CoVacU5) – an Investigator-Initiated Retrospective Cohort Study"

Supplementary Table 7 ........................................................................................ p.8-9

Supplementary Table 8 ........................................................................................ p.10-11

Supplementary Table 9 ........................................................................................ p.12-13

Supplementary Table 10 ......................................................................................p.14-15

Supplementary Table 11 ......................................................................................p.16

Supplementary Table 12 ...................................................................................... p.17.

Supplementary Table 13 ......................................................................................p.18-19

Supplementary Table 14 ......................................................................................p.20-21

Supplementary Table 15 ......................................................................................p.22-23

Supplementary Table 16 ......................................................................................p.24

Supplementary Table 17 ......................................................................................p.25

Suppl. Table 1: Comorbidities, n (%)

| Comorbidity | All N=7784 |
|---|---|
| None | 7100 (91.2) |
| Pulmonary diseases | 190 (2.4) |
| Malignant diseases | 7 (0.1) |
| Rheumatological diseases | 5 (0.1) |
| Gastroenterological diseases | 17 (0.2) |
| Carrdiological diseases | 166 (2.1) |
| Immunological diseases | 9 (0.1) |
| Trisomy 21 | 90 (1.2) |
| Other diseases | 305 (3.9) |

Suppl. Table 2: Post-vaccination symptoms after first BNT162b2 vaccine dose, n/N (%)

| | <12 months | | | | 12 to <24 months | | | | 24 to <60 months | | | |
|---|---|---|---|---|---|---|---|---|---|---|---|---|
| Symptoms | 3µg | 5µg | 10µg | p-value* | 3µg | 5µg | 10µg | p-value* | 3µg | 5µg | 10µg | p-value* |
| Local | 22/148 (14.9) | 23/170 (13.5) | 0/17 (0) | >0.999 | 110/515 (21.4) | 154/651 (23.7) | 29/97 (29.9) | >0.999 | 406/1094 (37.1) | 806/2281 (35.3) | 1023/2216 (46.2) | <0.001 |
| General | 17/148 (11.5) | 21/170 (12.4) | 1/17 (5.9) | >0.999 | 50/511 (9.8) | 99/650 (15.2) | 10/97 (10.3) | 0.1822 | 119/1090 (10.9) | 277/2278 (12.2) | 274/2208 (12.4) | >0.999 |
| Fever | 4/148 (2.7) | 6/170 (3.5) | 0/17 (0) | >0.999 | 19/511 (3.7) | 33/650 (5.1) | 5/97 (5.2) | >0.999 | 27/1090 (2.5) | 86/2278 (3.8) | 62/2208 (2.8) | 0.7391 |
| Musculoskeletal system | 2/148 (1.4) | 2/170 (1.2) | 0/17 (0) | >0.999 | 3/511 (0.6) | 11/649 (1.7) | 1/97 (1) | >0.999 | 19/1086 (1.7) | 65/2273 (2.9) | 90/2201 (4.1) | 0.0098 |
| Gastrointestinal | 3/148 (2) | 3/169 (1.8) | 0/17 (0) | >0.999 | 7/512 (1.4) | 14/647 (2.2) | 2/96 (2.1) | >0.999 | 16/1088 (1.5) | 42/2275 (1.8) | 48/2200 (2.2) | >0.999 |
| Otolaryngological | 0/148 (0) | 0/169 (0) | 0/17 (0) | - | 2/510 (0.4) | 9/644 (1.4) | 0/96 (0) | >0.999 | 13/1084 (1.2) | 23/2271 (1) | 25/2194 (1.1) | >0.999 |
| Pulmonary | 1/147 (0.7) | 0/169 (0) | 0/17 (0) | >0.999 | 8/511 (1.6) | 8/643 (1.2) | 1/96 (1) | >0.999 | 9/1085 (0.8) | 20/2269 (0.9) | 14/2195 (0.6) | >0.999 |
| Cardiovascular | 0/148 (0) | 0/169 (0) | 0/17 (0) | - | 1/510 (0.2) | 2/642 (0.3) | 0/96 (0) | >0.999 | 2/1086 (0.2) | 5/2268 (0.2) | 7/2197 (0.3) | >0.999 |
| Neurological | 0/147 (0) | 1/169 (0.6) | 0/17 (0) | >0.999 | 1/510 (0.2) | 3/642 (0.5) | 0/96 (0) | >0.999 | 8/1081 (0.7) | 19/2262 (0.8) | 30/2191 (1.4) | >0.999 |
| Psychological | 4/148 (2.7) | 2/169 (1.2) | 0/17 (0) | >0.999 | 7/511 (1.4) | 9/640 (1.4) | 1/96 (1) | >0.999 | 9/1083 (0.8) | 37/2264 (1.6) | 24/2190 (1.1) | >0.999 |
| Dermatological | 2/148 (1.4) | 1/169 (0.6) | 0/17 (0) | >0.999 | 16/508 (3.1) | 8/640 (1.3) | 0/96 (0) | 0.2655 | 15/1084 (1.4) | 31/2261 (1.4) | 42/2183 (1.9) | >0.999 |

Notes: *Adjusted for multiple testing by Bonferroni correction.

Suppl. Table 3: Post-vaccination symptoms after second BNT162b2 vaccine dose, n/N (%)

| | <12 months | | | | 12 to <24 months | | | | 24 to <60 months | | | |
|---|---|---|---|---|---|---|---|---|---|---|---|---|
| Symptoms | 3µg | 5µg | 10µg | p-value* | 3µg | 5µg | 10µg | p-value* | 3µg | 5µg | 10µg | p-value* |
| Local | 8/108 (7.4) | 26/162 (16) | 0/20 (0) | 0.2550 | 68/378 (18) | 145/628 (23.1) | 38/141 (27) | 0.5464 | 222/676 (32.8) | 640/1936 (33.1) | 1024/2522 (40.6) | <0.001 |
| General | 11/108 (10.2) | 17/162 (10.5) | 1/20 (5) | >0.999 | 45/373 (12.1) | 79/629 (12.6) | 19/141 (13.5) | >0.999 | 68/675 (10.1) | 197/1932 (10.2) | 262/2516 (10.4) | >0.999 |
| Fever | 2/108 (1.9) | 3/162 (1.9) | 0/20 (0) | >0.999 | 23/373 (6.2) | 46/629 (7.3) | 8/141 (5.7) | >0.999 | 21/675 (3.1) | 67/1932 (3.5) | 89/2516 (3.5) | >0.999 |
| Musculoskeletal system | 0/108 (0) | 2/162 (1.2) | 0/20 (0) | >0.999 | 4/373 (1.1) | 9/628 (1.4) | 1/141 (0.7) | >0.999 | 12/674 (1.8) | 52/1926 (2.7) | 96/2508 (3.8) | 0.1082 |
| Gastrointestinal | 2/108 (1.9) | 0/161 (0) | 0/20 (0) | >0.999 | 7/374 (1.9) | 18/626 (2.9) | 3/140 (2.1) | >0.999 | 11/674 (1.6) | 36/1929 (1.9) | 52/2505 (2.1) | >0.999 |
| Otolaryngological | 0/108 (0) | 0/161 (0) | 0/20 (0) | - | 1/374 (0.3) | 9/623 (1.4) | 2/140 (1.4) | >0.999 | 5/672 (0.7) | 24/1925 (1.2) | 23/2501 (0.9) | >0.999 |
| Pulmonary | 1/107 (0.9) | 0/161 (0) | 0/20 (0) | >0.999 | 3/374 (0.8) | 6/624 (1) | 2/140 (1.4) | >0.999 | 1/671 (0.1) | 10/1923 (0.5) | 19/2502 (0.8) | >0.999 |
| Cardiovascular | 1/108 (0.9) | 0/161 (0) | 0/20 (0) | >0.999 | 0/373 (0) | 2/623 (0.3) | 0/139 (0) | >0.999 | 0/671 (0) | 2/1925 (0.1) | 9/2503 (0.4) | 0.9144 |
| Neurological | 0/107 (0) | 1/161 (0.6) | 0/20 (0) | >0.999 | 1/373 (0.3) | 1/622 (0.2) | 0/140 (0) | >0.999 | 4/668 (0.6) | 14/1919 (0.7) | 30/2498 (1.2) | >0.999 |
| Psychological | 2/108 (1.9) | 2/161 (1.2) | 0/20 (0) | >0.999 | 3/374 (0.8) | 11/620 (1.8) | 0/140 (0) | >0.999 | 6/669 (0.9) | 28/1922 (1.5) | 22/2496 (0.9) | >0.999 |
| Dermatological | 0/108 (0) | 1/161 (0.6) | 0/20 (0) | >0.999 | 9/372 (2.4) | 14/620 (2.3) | 5/140 (3.6) | >0.999 | 11/671 (1.6) | 24/1916 (1.3) | 62/2492 (2.5) | 0.1147 |

Notes: *Adjusted for multiple testing by Bonferroni correction.

Suppl. Table 4: Post-vaccination symptoms after third BNT162b2 vaccine dose, n/N (%)

| | <12 months | | | | 12 to <24 months | | | | 24 to <60 months | | | |
|---|---|---|---|---|---|---|---|---|---|---|---|---|
| Symptoms | 3µg | 5µg | 10µg | p-value* | 3µg | 5µg | 10µg | p-value* | 3µg | 5µg | 10µg | p-value* |
| Local | 0/3 (0) | 3/11 (27.3) | 1/4 (25) | >0.999 | 1/15 (6.7) | 14/37 (37.8) | 11/46 (23.9) | 0.6606 | 11/34 (32.4) | 60/167 (35.9) | 192/475 (40.4) | >0.999 |
| General | 1/3 (33.3) | 2/11 (18.2) | 1/4 (25) | >0.999 | 0/15 (0) | 5/37 (13.5) | 7/46 (15.2) | >0.999 | 2/34 (5.9) | 17/167 (10.2) | 60/475 (12.6) | >0.999 |
| Fever | 0/3 (0) | 1/11 (9.1) | 0/4 (0) | >0.999 | 0/15 (0) | 2/37 (5.4) | 3/46 (6.5) | >0.999 | 0/34 (0) | 9/167 (5.4) | 26/475 (5.5) | >0.999 |
| Musculoskeletal system | 0/3 (0) | 0/11 (0) | 0/4 (0) | - | 0/15 (0) | 0/37 (0) | 0/46 (0) | - | 0/33 (0) | 6/167 (3.6) | 22/472 (4.7) | >0.999 |
| Gastrointestinal | 1/3 (33.3) | 1/11 (9.1) | 0/4 (0) | >0.999 | 0/15 (0) | 1/37 (2.7) | 1/46 (2.2) | >0.999 | 0/34 (0) | 2/167 (1.2) | 6/474 (1.3) | >0.999 |
| Otolaryngological | 0/3 (0) | 0/11 (0) | 0/4 (0) | - | 0/15 (0) | 0/37 (0) | 1/46 (2.2) | >0.999 | 2/34 (5.9) | 5/167 (3) | 1/474 (0.2) | 0.0064 |
| Pulmonary | 0/3 (0) | 1/11 (9.1) | 0/4 (0) | >0.999 | 0/15 (0) | 0/37 (0) | 0/46 (0) | - | 0/34 (0) | 1/167 (0.6) | 5/473 (1.1) | >0.999 |
| Cardiovascular | 0/3 (0) | 0/11 (0) | 0/4 (0) | - | 0/15 (0) | 0/37 (0) | 0/46 (0) | - | 0/34 (0) | 0/167 (0) | 1/472 (0.2) | >0.999 |
| Neurological | 0/3 (0) | 0/11 (0) | 0/4 (0) | - | 0/15 (0) | 0/37 (0) | 2/46 (4.3) | >0.999 | 1/34 (2.9) | 1/166 (0.6) | 6/471 (1.3) | >0.999 |
| Psychological | 0/3 (0) | 0/11 (0) | 0/4 (0) | - | 0/15 (0) | 0/37 (0) | 0/46 (0) | - | 0/34 (0) | 0/167 (0) | 2/470 (0.4) | >0.999 |
| Dermatological | 0/3 (0) | 1/11 (9.1) | 0/4 (0) | >0.999 | 0/14 (0) | 1/37 (2.7) | 0/46 (0) | >0.999 | 1/34 (2.9) | 3/167 (1.8) | 17/471 (3.6) | >0.999 |

Notes: *Adjusted for multiple testing by Bonferroni correction.

Suppl. Table 5: Post-vaccination symptoms/aftermath after any BNT162b2 vaccine dose, n/N (%)

| | <12 months | | | | 12 to <24 months | | | | 24 to <60 months | | | |
|---|---|---|---|---|---|---|---|---|---|---|---|---|
| | 3µg | 5µg | 10µg | p-value | 3µg | 5µg | 10µg | p-value | 3µg | 5µg | 10µg | p-value |
| Number absent days (mean,N) | 1/1 | 1/1 | -/0 | - | 2/18 | 3.5/21 | 1.5/4 | 0.2812 | 4.6/32 | 3/118 | 3/154 | 0.3457 |
| Susceptibility to infections | 1/100 (1) | 0/137 (0) | 0/13 (0) | 0.4709 | 3/337 (0.9) | 4/508 (0.8) | 0/79 (0) | 0.7090 | 5/461 (1.1) | 10/1532 (0.7) | 20/1898 (1.1) | 0.4210 |
| Threat rating scale (1-10), (mean,N) | 0.1/99 | 0.1/135 | 0/13 | 0.8365 | 0.1/332 | 0.2/499 | 0.1/79 | 0.2540 | 0.1/454 | 0.1/1520 | 0.1/1885 | 0.9182 |
| Threat rating scale=0 | 94/99 (94.9) | 131/135 (97) | 13/13 (100) | 0.5412 | 307/332 (92.5) | 457/499 (91.6) | 75/79 (94.9) | 0.5712 | 417/454 (91.9) | 1435/1520 (94.4) | 1757/1885 (93.2) | 0.1126 |
| Threat rating scale=1 | 3/99 (3) | 1/135 (0.7) | 0/13 (0) | 0.3490 | 21/332 (6.3) | 22/499 (4.4) | 3/79 (3.8) | 0.4044 | 29/454 (6.4) | 54/1520 (3.6) | 73/1885 (3.9) | 0.0233 |
| Threat rating scale=2 | 1/99 (1) | 1/135 (0.7) | 0/13 (0) | 0.9214 | 1/332 (0.3) | 13/499 (2.6) | 0/79 (0) | 0.0155 | 6/454 (1.3) | 15/1520 (1) | 38/1885 (2) | 0.0482 |
| Threat rating scale=3 | 1/99 (1) | 1/135 (0.7) | 0/13 (0) | 0.9214 | 2/332 (0.6) | 2/499 (0.4) | 1/79 (1.3) | 0.6185 | 1/454 (0.2) | 7/1520 (0.5) | 9/1885 (0.5) | 0.7503 |
| Threat rating scale=4 | 0/99 (0) | 0/135 (0) | 0/13 (0) | - | 0/332 (0) | 2/499 (0.4) | 0/79 (0) | 0.4380 | 1/454 (0.2) | 2/1520 (0.1) | 4/1885 (0.2) | 0.8416 |
| Threat rating scale=5 | 0/99 (0) | 0/135 (0) | 0/13 (0) | - | 1/332 (0.3) | 2/499 (0.4) | 0/79 (0) | 0.8410 | 0/454 (0) | 3/1520 (0.2) | 2/1885 (0.1) | 0.5463 |
| Threat rating scale=6 | 0/99 (0) | 0/135 (0) | 0/13 (0) | - | 0/332 (0) | 1/499 (0.2) | 0/79 (0) | 0.6621 | 0/454 (0) | 1/1520 (0.1) | 1/1885 (0.1) | 0.8637 |
| Threat rating scale=7 | 0/99 (0) | 1/135 (0.7) | 0/13 (0) | 0.6593 | 0/332 (0) | 0/499 (0) | 0/79 (0) | - | 0/454 (0) | 0/1520 (0) | 1/1885 (0.1) | 0.5923 |
| Threat rating scale=8 | 0/99 (0) | 0/135 (0) | 0/13 (0) | - | 0/332 (0) | 0/499 (0) | 0/79 (0) | - | 0/454 (0) | 0/1520 (0) | 0/1885 (0) | - |
| Threat rating scale=9 | 0/99 (0) | 0/135 (0) | 0/13 (0) | - | 0/332 (0) | 0/499 (0) | 0/79 (0) | - | 0/454 (0) | 1/1520 (0.1) | 0/1885 (0) | 0.4632 |
| Threat rating scale=10 | 0/99 (0) | 0/135 (0) | 0/13 (0) | - | 0/332 (0) | 0/499 (0) | 0/79 (0) | - | 0/454 (0) | 2/1520 (0.1) | 0/1885 (0) | 0.2145 |

Notes: *ANOVA

Suppl. Table 6: Local symptoms

| | Redness at the injection site | Swelling at the injection site | Pain at the injection site | Other local discomfort |
|---|---|---|---|---|
| After any vac. | 934/3363 (27.8) | 671/3363 (20) | 2789/3363 (82.9) | 52/3363 (1.5) |
| After 1st vac. | 791/3363 (23.5) | 530/3363 (15.8) | 2276/3363 (67.7) | 36/3363 (1.1) |
| After 2nd vac. | 632/3363 (18.8) | 448/3363 (13.3) | 1915/3363 (56.9) | 30/3363 (0.9) |
| Threat level (mean/N) | 0.2±0.6/919 | 0.2±0.7/658 | 0.1±0.6/2749 | 0.2±0.6/52 |
| *Dosage* | | | | |
| 3µg | 115/620 (18.5) | 75/462 (16.2) | 223/1838 (12.1) | 5/30 (16.7) |
| 5µg | 280/620 (45.2) | 198/462 (42.9) | 643/1838 (35) | 8/30 (26.7) |
| 10µg | 225/620 (36.3) | 189/462 (40.9) | 972/1838 (52.9) | 17/30 (56.7) |
| mixed/unknown | 314/934 (33.6) | 209/671 (31.1) | 951/2789 (34.1) | 22/52 (42.3) |
| *Age* | | | | |
| <12 months | 32/934 (3.4) | 21/671 (3.1) | 27/2789 (1) | 0/52 (0) |
| 12 to <24 months | 196/934 (21) | 129/671 (19.2) | 201/2789 (7.2) | 5/52 (9.6) |
| 24 to <60 months | 706/934 (75.6) | 521/671 (77.6) | 2561/2789 (91.8) | 47/52 (90.4) |
| *Beginning & duration* | | | | |
| Beginning (days after vaccination, mean/N) | 1.1±0.6/934 | 1±0.4/660 | 1±0.2/2764 | 1.1±0.7/52 |
| Duration (days, mean/N) | 2±1.8/915 | 2.4±3.9/651 | 1.6±1.1/2732 | 4.1±9.8/50 |
| > 90 days | 0/934 (0) | 0/671 (0) | 0/2789 (0) | 0/52 (0) |
| Ongoing (days, mean/N) | 0±0/0 | 0±0/0 | 0±0/0 | 93±0/1 |
| unknown | 6/934 (0.6) | 4/671 (0.6) | 12/2789 (0.4) | 0/52 (0) |
| *Aftermath* | | | | |
| ambulatory | 0/934 (0) | 0/671 (0) | 0/2789 (0) | 1/52 (1.9) |
| inpatient | 0/934 (0) | 0/671 (0) | 1/2789 (0) | 0/52 (0) |
| mortality | 0/934 (0) | 0/671 (0) | 0/2789 (0) | 0/52 (0) |
| other | 7/934 (0.7) | 8/671 (1.2) | 17/2789 (0.6) | 1/52 (1.9) |

Threat level from 0 (minimum) to 10 (maximum). Vac., vaccination. Ambulatory and inpatient refer to treatment requirement.

Suppl. Table 7: General symptoms

| | Fever | Chills | Hot flashes | Fatigue | Flu-like symptoms | Feeling of weakness | malaise/general feeling of illness | MIS-C | Other general complaints |
|---|---|---|---|---|---|---|---|---|---|
| After any vac. | 519/1505 (34.5) | 56/1505 (3.7) | 32/1505 (2.1) | 1043/1505 (69.3) | 131/1505 (8.7) | 140/1505 (9.3) | 342/1505 (22.7) | 0/1505 (0) | 100/1505 (6.6) |
| After 1st vac. | 267/1505 (17.7) | 28/1505 (1.9) | 19/1505 (1.3) | 766/1505 (50.9) | 74/1505 (4.9) | 100/1505 (6.6) | 215/1505 (14.3) | 0/1505 (0) | 66/1505 (4.4) |
| After 2nd vac. | 276/1505 (18.3) | 28/1505 (1.9) | 13/1505 (0.9) | 606/1505 (40.3) | 62/1505 (4.1) | 67/1505 (4.5) | 187/1505 (12.4) | 0/1505 (0) | 50/1505 (3.3) |
| Threat level (mean±SD/N) | 0.6±1.1/513 | 0.9±1.5/55 | 0.2±0.5/31 | 0.3±0.9/1028 | 1±1.3/128 | 0.8±1.4/136 | 0.6±1.1/337 | 0±0/0 | 0.8±1.2/98 |
| *Dosage* | | | | | | | | | |
| 3µg | 49/330 (14.8) | 4/37 (10.8) | 4/23 (17.4) | 104/689 (15.1) | 15/83 (18.1) | 12/93 (12.9) | 41/233 (17.6) | 0/0 (0) | 7/61 (11.5) |
| 5µg | 168/330 (50.9) | 12/37 (32.4) | 9/23 (39.1) | 296/689 (43) | 35/83 (42.2) | 30/93 (32.3) | 100/233 (42.9) | 0/0 (0) | 25/61 (41) |
| 10µg | 113/330 (34.2) | 21/37 (56.8) | 10/23 (43.5) | 289/689 (41.9) | 33/83 (39.8) | 51/93 (54.8) | 92/233 (39.5) | 0/0 (0) | 29/61 (47.5) |
| mixed/unknown | 189/519 (36.4) | 19/56 (33.9) | 9/32 (28.1) | 354/1043 (33.9) | 48/131 (36.6) | 47/140 (33.6) | 109/342 (31.9) | 0/0 (0) | 39/100 (39) |
| *Age* | | | | | | | | | |
| <12 months | 15/519 (2.9) | 0/56 (0) | 2/32 (6.3) | 37/1043 (3.5) | 2/131 (1.5) | 0/140 (0) | 14/342 (4.1) | 0/0 (0) | 3/100 (3) |
| 12 to <24 months | 125/519 (24.1) | 4/56 (7.1) | 5/32 (15.6) | 189/1043 (18.1) | 24/131 (18.3) | 17/140 (12.1) | 68/342 (19.9) | 0/0 (0) | 13/100 (13) |
| 24 to <60 months | 379/519 (73) | 52/56 (92.9) | 25/32 (78.1) | 817/1043 (78.3) | 105/131 (80.2) | 123/140 (87.9) | 260/342 (76) | 0/0 (0) | 84/100 (84) |
| *Beginning & duration* | | | | | | | | | |
| beginning (days after vaccination, mean±SD/N) | 1.1±0.4/519 | 1.1±0.3/56 | 1±0.2/31 | 1±0.2/1039 | 1.2±0.4/128 | 1.1±0.5/138 | 1±0.2/340 | 0±0/0 | 1.2±0.6/100 |
| duration (days, mean±SD/N) | 1.4±1.1/515 | 1.1±1/55 | 1±0.6/30 | 1.6±2.5/1026 | 2.9±2.4/128 | 2.3±2.9/138 | 1.9±1.7/337 | 0±0/0 | 2.8±4.5/99 |
| > 90 days | 0/519 (0) | 0/56 (0) | 0/32 (0) | 0/1043 (0) | 0/131 (0) | 0/140 (0) | 0/342 (0) | 0/0 (0) | 0/100 (0) |
| ongoing (days, mean±SD/N) | 0±0/0 | 0±0/0 | 0±0/0 | 0±0/0 | 0±0/0 | 0±0/0 | 0±0/0 | 0±0/0 | 86±0/1 |
| unknown | 0/519 (0) | 0/56 (0) | 0/32 (0) | 0/1043 (0) | 0/131 (0) | 0/140 (0) | 0/342 (0) | 0/0 (0) | 0/100 (0) |
| *Aftermath* | | | | | | | | | |
| ambulatory | 13/519 (2.5) | 1/56 (1.8) | 0/32 (0) | 4/1043 (0.4) | 9/131 (6.9) | 0/140 (0) | 4/342 (1.2) | 0/0 (0) | 4/100 (4) |
| inpatient | 0/519 (0) | 0/56 (0) | 0/32 (0) | 1/1043 (0.1) | 0/131 (0) | 1/140 (0.7) | 0/342 (0) | 0/0 (0) | 1/100 (1) |

| | | | | | | | | | |
|---|---|---|---|---|---|---|---|---|---|
| mortality | 0/519 (0) | 0/56 (0) | 0/32 (0) | 0/1043 (0) | 0/131 (0) | 0/140 (0) | 0/342 (0) | 0/0 (0) | 0/100 (0) |
| other | 1/519 (0.2) | 0/56 (0) | 0/32 (0) | 3/1043 (0.3) | 0/131 (0) | 0/140 (0) | 1/342 (0.3) | 0/0 (0) | 4/100 (4) |

Threat level from 0 (minimum) to 10 (maximum). Vac., vaccination. Ambulatory and inpatient refer to treatment requirement. MIS-C, multisystem inflammatory syndrome in children.

Suppl. Table 8: Musculoskeletal symptoms

| | muscle weakness | muscle pain | muscle twitching | neck/back pain | pain in the arms | pain in legs | joint pain | joint swelling | pain in limbs | other complaints |
|---|---|---|---|---|---|---|---|---|---|---|
| After any vac. | 11/319 (3.4) | 84/319 (26.3) | 1/319 (0.3) | 15/319 (4.7) | 179/319 (56.1) | 50/319 (15.7) | 12/319 (3.8) | 1/319 (0.3) | 29/319 (9.1) | 6/319 (1.9) |
| After 1st vac. | 9/319 (2.8) | 62/319 (19.4) | 0/319 (0) | 9/319 (2.8) | 133/319 (41.7) | 32/319 (10) | 8/319 (2.5) | 1/319 (0.3) | 16/319 (5) | 2/319 (0.6) |
| After 2nd vac. | 6/319 (1.9) | 53/319 (16.6) | 1/319 (0.3) | 10/319 (3.1) | 110/319 (34.5) | 26/319 (8.2) | 6/319 (1.9) | 0/319 (0) | 13/319 (4.1) | 4/319 (1.3) |
| Threat level (mean±SD/N) | 2.1±3.2/9 | 0.4±0.9/82 | 7±0/1 | 0.9±1.7/15 | 0.2±0.8/174 | 0.5±1.4/49 | 1.4±1.7/12 | 1±0/1 | 1.1±1.5/28 | 1.8±1.5/6 |
| *Dosage* | | | | | | | | | | |
| 3µg | 2/8 (25) | 3/48 (6.3) | 0/1 (0) | 0/9 (0) | 7/106 (6.6) | 5/34 (14.7) | 0/7 (0) | 0/1 (0) | 2/19 (10.5) | 1/3 (33.3) |
| 5µg | 3/8 (37.5) | 25/48 (52.1) | 1/1 (100) | 5/9 (55.6) | 29/106 (27.4) | 16/34 (47.1) | 2/7 (28.6) | 0/1 (0) | 4/19 (21.1) | 1/3 (33.3) |
| 10µg | 3/8 (37.5) | 20/48 (41.7) | 0/1 (0) | 4/9 (44.4) | 70/106 (66) | 13/34 (38.2) | 5/7 (71.4) | 1/1 (100) | 13/19 (68.4) | 1/3 (33.3) |
| mixed/unknown | 3/11 (27.3) | 36/84 (42.9) | 0/1 (0) | 6/15 (40) | 73/179 (40.8) | 16/50 (32) | 5/12 (41.7) | 0/1 (0) | 10/29 (34.5) | 3/6 (50) |
| *Age* | | | | | | | | | | |
| <12 months | 0/11 (0) | 3/84 (3.6) | 1/1 (100) | 0/15 (0) | 0/179 (0) | 1/50 (2) | 0/12 (0) | 0/1 (0) | 0/29 (0) | 0/6 (0) |
| 12 to <24 months | 1/11 (9.1) | 6/84 (7.1) | 0/1 (0) | 0/15 (0) | 10/179 (5.6) | 8/50 (16) | 0/12 (0) | 0/1 (0) | 2/29 (6.9) | 2/6 (33.3) |
| 24 to <60 months | 10/11 (90.9) | 75/84 (89.3) | 0/1 (0) | 15/15 (100) | 169/179 (94.4) | 41/50 (82) | 12/12 (100) | 1/1 (100) | 27/29 (93.1) | 4/6 (66.7) |
| *Beginning & duration* | | | | | | | | | | |
| beginning (days after vaccination, mean±SD/N) | 1.3±0.6/11 | 1±0.2/83 | 3±0/1 | 1±0/15 | 1±0.1/178 | 1.5±1.1/50 | 1.8±0.8/12 | 3±0/1 | 1±0/29 | 1.5±0.8/6 |
| duration (days, mean±SD/N) | 4.3±6.5/10 | 2.1±1.4/80 | 10±0/1 | 2.6±2.3/15 | 1.8±1.5/176 | 2.3±2.1/44 | 4±3.8/11 | 4±0/1 | 1.9±1.5/28 | 1.9±1.8/5 |
| > 90 days | 0/11 (0) | 0/84 (0) | 0/1 (0) | 0/15 (0) | 0/179 (0) | 0/50 (0) | 0/12 (0) | 0/1 (0) | 0/29 (0) | 0/6 (0) |
| ongoing (days, mean±SD/N) | 0±0/0 | 86±0/1 | 0±0/0 | 0±0/0 | 0±0/0 | 61±43.7/4 | 41±0/1 | 0±0/0 | 0±0/0 | 86±0/1 |
| unknown | 0/11 (0) | 0/84 (0) | 0/1 (0) | 0/15 (0) | 0/179 (0) | 0/50 (0) | 0/12 (0) | 0/1 (0) | 1/29 (3.4) | 0/6 (0) |
| *Aftermath* | | | | | | | | | | |
| ambulatory | 1/11 (9.1) | 1/84 (1.2) | 0/1 (0) | 1/15 (6.7) | 0/179 (0) | 2/50 (4) | 3/12 (25) | 1/1 (100) | 1/29 (3.4) | 0/6 (0) |
| inpatient | 1/11 (9.1) | 0/84 (0) | 1/1 (100) | 0/15 (0) | 0/179 (0) | 0/50 (0) | 0/12 (0) | 0/1 (0) | 0/29 (0) | 0/6 (0) |
| mortality | 0/11 (0) | 0/84 (0) | 0/1 (0) | 0/15 (0) | 0/179 (0) | 0/50 (0) | 0/12 (0) | 0/1 (0) | 0/29 (0) | 0/6 (0) |

| | | | | | | | | | | |
|---|---|---|---|---|---|---|---|---|---|---|
| other | 0/11 (0) | 0/84 (0) | 0/1 (0) | 0/15 (0) | 1/179 (0.6) | 1/50 (2) | 1/12 (8.3) | 0/1 (0) | 0/29 (0) | 0/6 (0) |

Threat level from 0 (minimum) to 10 (maximum). Vac., vaccination. Ambulatory and inpatient refer to treatment requirement.

Suppl. Table 9: Gastrointestinal symptoms

| | abdominal pain | nausea/vomiting | constipation | diarrhea | other stool changes | unwanted weight loss | unwanted weight gain | other gastrointestinal complaints |
|---|---|---|---|---|---|---|---|---|
| After any vac. | 61/259 (23.6) | 108/259 (41.7) | 14/259 (5.4) | 82/259 (31.7) | 45/259 (17.4) | 2/259 (0.8) | 0/259 (0) | 7/259 (2.7) |
| After 1st vac. | 31/259 (12) | 60/259 (23.2) | 6/259 (2.3) | 50/259 (19.3) | 25/259 (9.7) | 0/259 (0) | 0/259 (0) | 0/259 (0) |
| After 2nd vac. | 29/259 (11.2) | 50/259 (19.3) | 10/259 (3.9) | 47/259 (18.1) | 32/259 (12.4) | 2/259 (0.8) | 0/259 (0) | 5/259 (1.9) |
| Threat level (mean±SD/N) | 1.4±1.5/60 | 0.9±1.3/106 | 0.8±1.3/14 | 0.9±1.4/82 | 0.5±0.9/44 | 4±0/2 | 0±0/0 | 2±1.6/7 |
| *Dosage* | | | | | | | | |
| 3µg | 7/39 (17.9) | 8/73 (11) | 3/8 (37.5) | 4/50 (8) | 7/31 (22.6) | 0/2 (0) | 0/0 (0) | 1/4 (25) |
| 5µg | 10/39 (25.6) | 35/73 (47.9) | 3/8 (37.5) | 24/50 (48) | 12/31 (38.7) | 2/2 (100) | 0/0 (0) | 2/4 (50) |
| 10µg | 22/39 (56.4) | 30/73 (41.1) | 2/8 (25) | 22/50 (44) | 12/31 (38.7) | 0/2 (0) | 0/0 (0) | 1/4 (25) |
| mixed/unknown | 22/61 (36.1) | 35/108 (32.4) | 6/14 (42.9) | 32/82 (39) | 14/45 (31.1) | 0/2 (0) | 0/0 (0) | 3/7 (42.9) |
| *Age* | | | | | | | | |
| <12 months | 1/61 (1.6) | 1/108 (0.9) | 0/14 (0) | 4/82 (4.9) | 4/45 (8.9) | 0/2 (0) | 0/0 (0) | 1/7 (14.3) |
| 12 to <24 months | 3/61 (4.9) | 18/108 (16.7) | 3/14 (21.4) | 17/82 (20.7) | 9/45 (20) | 2/2 (100) | 0/0 (0) | 0/7 (0) |
| 24 to <60 months | 57/61 (93.4) | 89/108 (82.4) | 11/14 (78.6) | 61/82 (74.4) | 32/45 (71.1) | 0/2 (0) | 0/0 (0) | 6/7 (85.7) |
| *Beginning & duration* | | | | | | | | |
| beginning (days after vaccination, mean±SD/N) | 1.6±1/60 | 1.2±0.6/108 | 2.1±1.8/14 | 1.5±0.8/82 | 1.3±0.9/45 | 1±0/2 | 0±0/0 | 2.2±1.6/6 |
| duration (days, mean±SD/N) | 4.6±9.4/53 | 1.3±1.6/108 | 11.2±10.7/13 | 4.3±7.8/81 | 4.3±7/45 | 4±1.4/2 | 0±0/0 | 2±1/3 |
| > 90 days | 0/61 (0) | 0/108 (0) | 0/14 (0) | 0/82 (0) | 0/45 (0) | 0/2 (0) | 0/0 (0) | 0/7 (0) |
| ongoing (days, mean±SD/N) | 45.7±38.2/3 | 0±0/0 | 55±0/1 | 0±0/0 | 0±0/0 | 0±0/0 | 0±0/0 | 79±53.7/2 |
| unknown | 2/61 (3.3) | 0/108 (0) | 0/14 (0) | 0/82 (0) | 0/45 (0) | 0/2 (0) | 0/0 (0) | 0/7 (0) |
| *Aftermath* | | | | | | | | |
| ambulatory | 4/61 (6.6) | 1/108 (0.9) | 1/14 (7.1) | 7/82 (8.5) | 2/45 (4.4) | 0/2 (0) | 0/0 (0) | 1/7 (14.3) |
| inpatient | 1/61 (1.6) | 0/108 (0) | 0/14 (0) | 0/82 (0) | 0/45 (0) | 0/2 (0) | 0/0 (0) | 0/7 (0) |

| | | | | | | | | |
|---|---|---|---|---|---|---|---|---|
| mortality | 0/61 (0) | 0/108 (0) | 0/14 (0) | 0/82 (0) | 0/45 (0) | 0/2 (0) | 0/0 (0) | 0/7 (0) |
| other | 0/61 (0) | 2/108 (1.9) | 0/14 (0) | 1/82 (1.2) | 0/45 (0) | 0/2 (0) | 0/0 (0) | 1/7 (14.3) |

Threat level from 0 (minimum) to 10 (maximum). Vac., vaccination. Ambulatory and inpatient refer to treatment requirement.

Suppl. Table 10: Otolaryngological symptoms

| | Nosebleed | redness of the oral mucosa | swelling of the tongue | swelling of the lips | discomfort in the mouth | toothache | bleeding of the gums | tightness in the throat | sore throat | earache | hoarseness | facial swelling | swollen lymph nodes | painful lymph nodes | olfactory disorder | taste disorder | other complaints |
|---|---|---|---|---|---|---|---|---|---|---|---|---|---|---|---|---|---|
| After any vac. | 17/132 (12.9) | 4/132 (3) | 0/132 (0) | 0/132 (0) | 1/132 (0.8) | 6/132 (4.5) | 0/132 (0) | 2/132 (1.5) | 20/132 (15.2) | 25/132 (18.9) | 13/132 (9.8) | 2/132 (1.5) | 44/132 (33.3) | 5/132 (3.8) | 1/132 (0.8) | 2/132 (1.5) | 18/132 (13.6) |
| After 1st vac. | 10/132 (7.6) | 3/132 (2.3) | 0/132 (0) | 0/132 (0) | 0/132 (0) | 2/132 (1.5) | 0/132 (0) | 2/132 (1.5) | 12/132 (9.1) | 12/132 (9.1) | 5/132 (3.8) | 0/132 (0) | 29/132 (22) | 3/132 (2.3) | 1/132 (0.8) | 1/132 (0.8) | 11/132 (8.3) |
| After 2nd vac. | 9/132 (6.8) | 2/132 (1.5) | 0/132 (0) | 0/132 (0) | 1/132 (0.8) | 5/132 (3.8) | 0/132 (0) | 0/132 (0) | 9/132 (6.8) | 13/132 (9.8) | 6/132 (4.5) | 2/132 (1.5) | 28/132 (21.2) | 2/132 (1.5) | 0/132 (0) | 1/132 (0.8) | 7/132 (5.3) |
| Threat level (mean±SD/N) | 0.8±1.3/17 | 0.8±1/4 | 0±0/0 | 0±0/0 | 2±0/1 | 1±2/6 | 0±0/0 | 3±1.4/2 | 1.7±1.8/20 | 1.2±1.4/25 | 1.1±1.7/13 | 1.5±2.1/2 | 0.6±1.2/42 | 0.2±0.4/5 | 1±0/1 | 1.5±0.7/2 | 0.6±0.8/18 |
| *Dosage* | | | | | | | | | | | | | | | | | |
| 3µg | 0/8 (0) | 0/3 (0) | 0/0 (0) | 0/0 (0) | 0/1 (0) | 0/4 (0) | 0/0 (0) | 0/2 (0) | 3/15 (20) | 3/14 (21.4) | 0/9 (0) | 0/1 (0) | 2/27 (7.4) | 0/4 (0) | 0/0 (0) | 0/0 (0) | 2/12 (16.7) |
| 5µg | 4/8 (50) | 2/3 (66.7) | 0/0 (0) | 0/0 (0) | 0/1 (0) | 3/4 (75) | 0/0 (0) | 1/2 (50) | 6/15 (40) | 5/14 (35.7) | 6/9 (66.7) | 1/1 (100) | 13/27 (48.1) | 0/4 (0) | 0/0 (0) | 0/0 (0) | 6/12 (50) |
| 10µg | 4/8 (50) | 1/3 (33.3) | 0/0 (0) | 0/0 (0) | 1/1 (100) | 1/4 (25) | 0/0 (0) | 1/2 (50) | 6/15 (40) | 6/14 (42.9) | 3/9 (33.3) | 0/1 (0) | 12/27 (44.4) | 4/4 (100) | 0/0 (0) | 0/0 (0) | 4/12 (33.3) |
| mixed/unknown | 9/17 (52.9) | 1/4 (25) | 0/0 (0) | 0/0 (0) | 0/1 (0) | 2/6 (33.3) | 0/0 (0) | 0/2 (0) | 5/20 (25) | 11/25 (44) | 4/13 (30.8) | 1/2 (50) | 17/44 (38.6) | 1/5 (20) | 1/1 (100) | 2/2 (100) | 6/18 (33.3) |
| *Age* | | | | | | | | | | | | | | | | | |
| <12 months | 0/17 (0) | 0/4 (0) | 0/0 (0) | 0/0 (0) | 0/1 (0) | 0/6 (0) | 0/0 (0) | 0/2 (0) | 0/20 (0) | 0/25 (0) | 0/13 (0) | 0/2 (0) | 0/44 (0) | 0/5 (0) | 0/1 (0) | 0/2 (0) | 1/18 (5.6) |
| 12 to <24 months | 1/17 (5.9) | 0/4 (0) | 0/0 (0) | 0/0 (0) | 0/1 (0) | 3/6 (50) | 0/0 (0) | 0/2 (0) | 4/20 (20) | 4/25 (16) | 3/13 (23.1) | 0/2 (0) | 7/44 (15.9) | 0/5 (0) | 0/1 (0) | 0/2 (0) | 2/18 (11.1) |
| 24 to <60 months | 16/17 (94.1) | 4/4 (100) | 0/0 (0) | 0/0 (0) | 1/1 (100) | 3/6 (50) | 0/0 (0) | 2/2 (100) | 16/20 (80) | 21/25 (84) | 10/13 (76.9) | 2/2 (100) | 37/44 (84.1) | 5/5 (100) | 1/1 (100) | 2/2 (100) | 15/18 (83.3) |
| *beginning & duration* | | | | | | | | | | | | | | | | | |
| beginning (days after vaccination, mean±SD/N) | 2.2±1.6/17 | 1.5±0.6/4 | 0±0/0 | 0±0/0 | 1±0/1 | 2.2±1.6/6 | 0±0/0 | 1±0/2 | 1.5±0.7/20 | 1.6±0.9/25 | 1.4±0.7/12 | 1±0/2 | 1.6±1.2/44 | 1.2±0.4/5 | 1±0/1 | 2±0/2 | 2.6±2/18 |
| duration (days, mean±SD/N) | 1.9±2.2/17 | 4.5±1.7/4 | 0±0/0 | 0±0/0 | 1±0/1 | 5.2±1.8/6 | 0±0/0 | 3±0/2 | 3.4±2/19 | 2.3±1.6/23 | 2.3±1.2/11 | 1.3±1.1/2 | 5.3±9/43 | 2±0.8/4 | 16±0/1 | 9±7.1/2 | 12±23.2/13 |
| > 90 days | 0/17 (0) | 0/4 (0) | 0/0 (0) | 0/0 (0) | 0/1 (0) | 0/6 (0) | 0/0 (0) | 0/2 (0) | 0/20 (0) | 0/25 (0) | 0/13 (0) | 0/2 (0) | 0/44 (0) | 0/5 (0) | 0/1 (0) | 0/2 (0) | 0/18 (0) |
| ongoing (days, mean±SD/N) | 0±0/0 | 0±0/0 | 0±0/0 | 0±0/0 | 0±0/0 | 0±0/0 | 0±0/0 | 0±0/0 | 0±0/0 | 41±0/1 | 0±0/0 | 0±0/0 | 0±0/0 | 0±0/0 | 0±0/0 | 0±0/0 | 57±23.7/4 |

| | | | | | | | | | | | | | | | | | |
|---|---|---|---|---|---|---|---|---|---|---|---|---|---|---|---|---|---|
| unknown | 0/17 (0) | 0/4 (0) | 0/0 (0) | 0/0 (0) | 0/1 (0) | 0/6 (0) | 0/0 (0) | 0/2 (0) | 0/20 (0) | 0/25 (0) | 0/13 (0) | 0/2 (0) | 0/44 (0) | 0/5 (0) | 0/1 (0) | 0/2 (0) | 1/18 (5.6) |
| *Aftermath* | | | | | | | | | | | | | | | | | |
| ambulatory | 0/17 (0) | 0/4 (0) | 0/0 (0) | 0/0 (0) | 0/1 (0) | 0/6 (0) | 0/0 (0) | 0/2 (0) | 5/20 (25) | 10/25 (40) | 1/13 (7.7) | 0/2 (0) | 0/44 (0) | 0/5 (0) | 0/1 (0) | 0/2 (0) | 6/18 (33.3) |
| inpatient | 0/17 (0) | 0/4 (0) | 0/0 (0) | 0/0 (0) | 0/1 (0) | 0/6 (0) | 0/0 (0) | 0/2 (0) | 0/20 (0) | 0/25 (0) | 0/13 (0) | 0/2 (0) | 1/44 (2.3) | 0/5 (0) | 0/1 (0) | 0/2 (0) | 0/18 (0) |
| mortality | 0/17 (0) | 0/4 (0) | 0/0 (0) | 0/0 (0) | 0/1 (0) | 0/6 (0) | 0/0 (0) | 0/2 (0) | 0/20 (0) | 0/25 (0) | 0/13 (0) | 0/2 (0) | 0/44 (0) | 0/5 (0) | 0/1 (0) | 0/2 (0) | 0/18 (0) |
| other | 0/17 (0) | 0/4 (0) | 0/0 (0) | 0/0 (0) | 0/1 (0) | 0/6 (0) | 0/0 (0) | 0/2 (0) | 1/20 (5) | 1/25 (4) | 0/13 (0) | 1/2 (50) | 0/44 (0) | 0/5 (0) | 0/1 (0) | 0/2 (0) | 2/18 (11.1) |

Threat level from 0 (minimum) to 10 (maximum). Vac., vaccination. Ambulatory and inpatient refer to treatment requirement.

Suppl. Table 11: Pulmonary symptoms

| | cough | irregular breathing | rapid breathing | shortness of breath on exertion | dyspnea at rest | other breathing problems |
|---|---|---|---|---|---|---|
| After any vac. | 88/104 (84.6) | 2/104 (1.9) | 7/104 (6.7) | 6/104 (5.8) | 7/104 (6.7) | 13/104 (12.5) |
| After 1st vac. | 55/104 (52.9) | 2/104 (1.9) | 4/104 (3.8) | 5/104 (4.8) | 5/104 (4.8) | 8/104 (7.7) |
| After 2nd vac. | 36/104 (34.6) | 0/104 (0) | 3/104 (2.9) | 1/104 (1) | 1/104 (1) | 5/104 (4.8) |
| Threat level (mean/N) | 1.2±1.6/86 | 0.5±0.7/2 | 2.1±1.6/7 | 2.2±2.4/6 | 2.1±2.5/7 | 2.8±2.1/13 |
| *Dosage* | | | | | | |
| 3µg | 8/57 (14) | 0/2 (0) | 1/3 (33.3) | 0/3 (0) | 0/5 (0) | 1/9 (11.1) |
| 5µg | 25/57 (43.9) | 1/2 (50) | 1/3 (33.3) | 2/3 (66.7) | 1/5 (20) | 5/9 (55.6) |
| 10µg | 24/57 (42.1) | 1/2 (50) | 1/3 (33.3) | 1/3 (33.3) | 4/5 (80) | 3/9 (33.3) |
| mixed/unknown | 31/88 (35.2) | 0/2 (0) | 4/7 (57.1) | 3/6 (50) | 2/7 (28.6) | 4/13 (30.8) |
| *Age* | | | | | | |
| <12 months | 2/88 (2.3) | 0/2 (0) | 0/7 (0) | 0/6 (0) | 1/7 (14.3) | 0/13 (0) |
| 12 to <24 months | 19/88 (21.6) | 0/2 (0) | 1/7 (14.3) | 2/6 (33.3) | 0/7 (0) | 3/13 (23.1) |
| 24 to <60 months | 67/88 (76.1) | 2/2 (100) | 6/7 (85.7) | 4/6 (66.7) | 6/7 (85.7) | 10/13 (76.9) |
| *Beginning &duration* | | | | | | |
| beginning (days after vaccination, mean/N) | 1.5±0.9/86 | 2±0/2 | 1.1±0.4/7 | 2.3±2/6 | 1.7±0.8/7 | 1.2±0.4/13 |
| duration (days, mean/N) | 5.7±4.2/82 | 8±8.5/2 | 7.9±10.7/7 | 8.8±7.3/5 | 3.8±3.6/6 | 4±3.1/12 |
| > 90 days | 0/88 (0) | 0/2 (0) | 0/7 (0) | 0/6 (0) | 0/7 (0) | 0/13 (0) |
| ongoing (days, mean/N) | 18±21.2/2 | 0±0/0 | 0±0/0 | 0±0/0 | 0±0/0 | 3±0/1 |
| unknown | 0/88 (0) | 0/2 (0) | 0/7 (0) | 0/6 (0) | 0/7 (0) | 0/13 (0) |
| *Aftermath* | | | | | | |
| ambulatory | 9/88 (10.2) | 0/2 (0) | 2/7 (28.6) | 1/6 (16.7) | 0/7 (0) | 4/13 (30.8) |
| inpatient | 2/88 (2.3) | 1/2 (50) | 2/7 (28.6) | 2/6 (33.3) | 3/7 (42.9) | 3/13 (23.1) |
| mortality | 0/88 (0) | 0/2 (0) | 0/7 (0) | 0/6 (0) | 0/7 (0) | 0/13 (0) |
| other | 2/88 (2.3) | 0/2 (0) | 0/7 (0) | 0/6 (0) | 1/7 (14.3) | 2/13 (15.4) |

Threat level from 0 (minimum) to 10 (maximum). Vac., vaccination. Ambulatory and inpatient refer to treatment requirement.

Suppl. Table 12: Cardiovascular symptoms

| | heart collapse | tachycardia | heart pain | chest tightness | cold hands and feet | other |
|---|---|---|---|---|---|---|
| After any vac. | 1/32 (3.1) | 11/32 (34.4) | 5/32 (15.6) | 1/32 (3.1) | 12/32 (37.5) | 2/32 (6.3) |
| After 1st vac. | 0/32 (0) | 3/32 (9.4) | 3/32 (9.4) | 1/32 (3.1) | 10/32 (31.3) | 0/32 (0) |
| After 2nd vac. | 1/32 (3.1) | 8/32 (25) | 3/32 (9.4) | 0/32 (0) | 2/32 (6.3) | 2/32 (6.3) |
| Threat level (mean/N) | 6±0/1 | 2.3±2.7/11 | 0.6±1.3/5 | 3±0/1 | 1.3±2.2/12 | 3±1.4/2 |
| *Dosage* | | | | | | |
| 3µg | 0/1 (0) | 1/9 (11.1) | 1/5 (20) | 0/0 (0) | 1/9 (11.1) | 0/2 (0) |
| 5µg | 0/1 (0) | 3/9 (33.3) | 1/5 (20) | 0/0 (0) | 5/9 (55.6) | 0/2 (0) |
| 10µg | 1/1 (100) | 5/9 (55.6) | 3/5 (60) | 0/0 (0) | 3/9 (33.3) | 2/2 (100) |
| mixed/unknown | 0/1 (0) | 2/11 (18.2) | 0/5 (0) | 1/1 (100) | 3/12 (25) | 0/2 (0) |
| *Age* | | | | | | |
| <12 months | 0/1 (0) | 1/11 (9.1) | 0/5 (0) | 0/1 (0) | 0/12 (0) | 0/2 (0) |
| 12 to <24 months | 0/1 (0) | 0/11 (0) | 1/5 (20) | 0/1 (0) | 3/12 (25) | 0/2 (0) |
| 24 to <60 months | 1/1 (100) | 10/11 (90.9) | 4/5 (80) | 1/1 (100) | 9/12 (75) | 2/2 (100) |
| *Beginning & duration* | | | | | | |
| beginning (days after vaccination, mean/N) | 3±0/1 | 1.5±0.9/11 | 1.8±0.4/5 | 1±0/1 | 1.3±0.6/12 | 1.5±0.7/2 |
| duration (days, mean/N) | 0.5±0/1 | 2.7±4/10 | 3.5±2.5/5 | 0.5±0/1 | 3.1±2.4/10 | 0.8±0.4/2 |
| > 90 days | 0/1 (0) | 0/11 (0) | 0/5 (0) | 0/1 (0) | 0/12 (0) | 0/2 (0) |
| ongoing (days, mean/N) | 0±0/0 | 32±0/1 | 0±0/0 | 0±0/0 | 0±0/0 | 0±0/0 |
| unknown | 0/1 (0) | 0/11 (0) | 0/5 (0) | 0/1 (0) | 0/12 (0) | 0/2 (0) |
| *Aftermath* | | | | | | |
| ambulatory | 0/1 (0) | 3/11 (27.3) | 0/5 (0) | 0/1 (0) | 0/12 (0) | 0/2 (0) |
| inpatient | 1/1 (100) | 1/11 (9.1) | 0/5 (0) | 0/1 (0) | 0/12 (0) | 0/2 (0) |
| mortality | 0/1 (0) | 0/11 (0) | 0/5 (0) | 0/1 (0) | 0/12 (0) | 0/2 (0) |
| other | 0/1 (0) | 0/11 (0) | 0/5 (0) | 0/1 (0) | 0/12 (0) | 0/2 (0) |

Threat level from 0 (minimum) to 10 (maximum). Vac., vaccination. Ambulatory and inpatient refer to treatment requirement

Suppl. Table 13:Neurological symptoms

| | headache | dizziness | sensory disturbances | locomotor disorders | loss of consciousnes s | shooting pain | seizure | paralysis of facial muscles | other complaints |
|---|---|---|---|---|---|---|---|---|---|
| After any vac. | 101/121 (83.5) | 12/121 (9.9) | 1/121 (0.8) | 5/121 (4.1) | 1/121 (0.8) | 1/121 (0.8) | 1/121 (0.8) | 0/121 (0) | 3/121 (2.5) |
| After 1st vac. | 54/121 (44.6) | 6/121 (5) | 1/121 (0.8) | 3/121 (2.5) | 1/121 (0.8) | 0/121 (0) | 0/121 (0) | 0/121 (0) | 1/121 (0.8) |
| After 2nd vac. | 44/121 (36.4) | 4/121 (3.3) | 0/121 (0) | 3/121 (2.5) | 0/121 (0) | 1/121 (0.8) | 1/121 (0.8) | 0/121 (0) | 3/121 (2.5) |
| Threat level (mean±SD/N) | 0.5±0.9/101 | 2.2±3/12 | 1±0/1 | 2.2±2.3/5 | 9±0/1 | 1±0/1 | 1±0/1 | 0±0/0 | 4±3/3 |
| *Dosage* | | | | | | | | | |
| 3µg | 9/64 (14.1) | 1/10 (10) | 0/1 (0) | 0/4 (0) | 0/1 (0) | 0/1 (0) | 0/1 (0) | 0/0 (0) | 0/1 (0) |
| 5µg | 16/64 (25) | 5/10 (50) | 0/1 (0) | 3/4 (75) | 1/1 (100) | 0/1 (0) | 1/1 (100) | 0/0 (0) | 1/1 (100) |
| 10µg | 39/64 (60.9) | 4/10 (40) | 1/1 (100) | 1/4 (25) | 0/1 (0) | 1/1 (100) | 0/1 (0) | 0/0 (0) | 0/1 (0) |
| mixed/unknown | 37/101 (36.6) | 2/12 (16.7) | 0/1 (0) | 1/5 (20) | 0/1 (0) | 0/1 (0) | 0/1 (0) | 0/0 (0) | 2/3 (66.7) |
| *Age* | | | | | | | | | |
| <12 months | 1/101 (1) | 0/12 (0) | 0/1 (0) | 0/5 (0) | 0/1 (0) | 0/1 (0) | 0/1 (0) | 0/0 (0) | 1/3 (33.3) |
| 12 to <24 months | 6/101 (5.9) | 1/12 (8.3) | 0/1 (0) | 2/5 (40) | 0/1 (0) | 0/1 (0) | 0/1 (0) | 0/0 (0) | 0/3 (0) |
| 24 to <60 months | 94/101 (93.1) | 11/12 (91.7) | 1/1 (100) | 3/5 (60) | 1/1 (100) | 1/1 (100) | 1/1 (100) | 0/0 (0) | 2/3 (66.7) |
| *Beginning & duration* | | | | | | | | | |
| beginning (days after vaccination, mean±SD/N) | 1.1±0.5/95 | 2.2±1.9/12 | 3±0/1 | 2.4±2.2/5 | 3±0/1 | 5±0/1 | 1±0/1 | 0±0/0 | 2.3±1.2/3 |
| duration (days, mean±SD/N) | 1.6±2.1/94 | 2.5±3.4/11 | 0±0/0 | 2.3±2.8/5 | 1±0/1 | 6±0/1 | 0.5±0/1 | 0±0/0 | 21±0/1 |
| > 90 days | 0/101 (0) | 0/12 (0) | 1/1 (100) | 0/5 (0) | 0/1 (0) | 0/1 (0) | 0/1 (0) | 0/0 (0) | 0/3 (0) |
| ongoing (days, mean±SD/N) | 66±0/1 | 0±0/0 | 0±0/0 | 0±0/0 | 0±0/0 | 0±0/0 | 0±0/0 | 0±0/0 | 86±0/1 |
| unknown | 0/101 (0) | 0/12 (0) | 0/1 (0) | 0/5 (0) | 0/1 (0) | 0/1 (0) | 0/1 (0) | 0/0 (0) | 0/3 (0) |
| *Aftermath* | | | | | | | | | |
| ambulatory | 0/101 (0) | 1/12 (8.3) | 0/1 (0) | 0/5 (0) | 0/1 (0) | 0/1 (0) | 0/1 (0) | 0/0 (0) | 0/3 (0) |
| inpatient | 1/101 (1) | 2/12 (16.7) | 0/1 (0) | 1/5 (20) | 1/1 (100) | 0/1 (0) | 0/1 (0) | 0/0 (0) | 1/3 (33.3) |

| | | | | | | | | | |
|---|---|---|---|---|---|---|---|---|---|
| mortality | 0/101 (0) | 0/12 (0) | 0/1 (0) | 0/5 (0) | 0/1 (0) | 0/1 (0) | 0/1 (0) | 0/0 (0) | 0/3 (0) |
| other | 1/101 (1) | 0/12 (0) | 0/1 (0) | 0/5 (0) | 0/1 (0) | 0/1 (0) | 1/1 (100) | 0/0 (0) | 0/3 (0) |

Threat level from 0 (minimum) to 10 (maximum). Vac., vaccination. Ambulatory and inpatient refer to treatment requirement.

.

Suppl. Table 14: Psychological symptoms

| | concentration disorders | memory disorder | sleep disorders | aggressive behavior | anxiousness | hyperactivity | timid behavior | sadness/ depression | mood swings | altered behavior or other complaints |
|---|---|---|---|---|---|---|---|---|---|---|
| After any vac. | 4/136 (2.9) | 1/136 (0.7) | 40/136 (29.4) | 10/136 (7.4) | 5/136 (3.7) | 41/136 (30.1) | 12/136 (8.8) | 7/136 (5.1) | 30/136 (22.1) | 16/136 (11.8) |
| After 1st vac. | 4/136 (2.9) | 0/136 (0) | 30/136 (22.1) | 7/136 (5.1) | 3/136 (2.2) | 36/136 (26.5) | 7/136 (5.1) | 3/136 (2.2) | 19/136 (14) | 11/136 (8.1) |
| After 2nd vac. | 2/136 (1.5) | 0/136 (0) | 26/136 (19.1) | 7/136 (5.1) | 5/136 (3.7) | 23/136 (16.9) | 9/136 (6.6) | 3/136 (2.2) | 18/136 (13.2) | 8/136 (5.9) |
| Threat level (mean±SD/N) | 0±0/4 | 1±0/1 | 0.3±0.6/40 | 1±1.2/10 | 1.4±3.1/5 | 0.2±0.7/41 | 0.8±1.3/12 | 2.1±3/7 | 0.8±1.5/30 | 0.8±1.1/16 |
| *Dosage* | | | | | | | | | | |
| 3µg | 0/3 (0) | 0/1 (0) | 3/27 (11.1) | 2/7 (28.6) | 0/3 (0) | 5/30 (16.7) | 1/7 (14.3) | 0/5 (0) | 3/19 (15.8) | 0/9 (0) |
| 5µg | 2/3 (66.7) | 0/1 (0) | 16/27 (59.3) | 3/7 (42.9) | 2/3 (66.7) | 16/30 (53.3) | 4/7 (57.1) | 2/5 (40) | 11/19 (57.9) | 8/9 (88.9) |
| 10µg | 1/3 (33.3) | 1/1 (100) | 8/27 (29.6) | 2/7 (28.6) | 1/3 (33.3) | 9/30 (30) | 2/7 (28.6) | 3/5 (60) | 5/19 (26.3) | 1/9 (11.1) |
| mixed/unknown | 1/4 (25) | 0/1 (0) | 13/40 (32.5) | 3/10 (30) | 2/5 (40) | 11/41 (26.8) | 5/12 (41.7) | 2/7 (28.6) | 11/30 (36.7) | 7/16 (43.8) |
| *Age* | | | | | | | | | | |
| <12 months | 0/4 (0) | 0/1 (0) | 4/40 (10) | 0/10 (0) | 1/5 (20) | 2/41 (4.9) | 0/12 (0) | 1/7 (14.3) | 2/30 (6.7) | 1/16 (6.3) |
| 12 to <24 months | 0/4 (0) | 0/1 (0) | 14/40 (35) | 1/10 (10) | 0/5 (0) | 5/41 (12.2) | 1/12 (8.3) | 0/7 (0) | 4/30 (13.3) | 3/16 (18.8) |
| 24 to <60 months | 4/4 (100) | 1/1 (100) | 22/40 (55) | 9/10 (90) | 4/5 (80) | 34/41 (82.9) | 11/12 (91.7) | 6/7 (85.7) | 24/30 (80) | 12/16 (75) |
| *Beginning & duration* | | | | | | | | | | |
| beginning (days after vaccination, mean±SD/N) | 1.5±1/4 | 3±0/1 | 1.1±0.4/40 | 2±1.4/10 | 1.8±1.1/5 | 1±0.3/41 | 1.7±1.2/12 | 2.8±2.2/6 | 1.2±0.6/30 | 1.4±0.7/16 |
| duration (days, mean±SD/N) | 4.1±3.4/4 | 7±0/1 | 2.5±2.7/38 | 6.1±7/8 | 4.7±4.7/3 | 1.8±2.6/38 | 5.9±9/8 | 4.2±3.2/3 | 2.5±3/25 | 6.1±8/15 |
| > 90 days | 0/4 (0) | 0/1 (0) | 0/40 (0) | 0/10 (0) | 0/5 (0) | 0/41 (0) | 0/12 (0) | 0/7 (0) | 0/30 (0) | 0/16 (0) |
| ongoing (days, mean±SD/N) | 0±0/0 | 0±0/0 | 0±0/1 | 48.5±27.6/2 | 68±0/1 | 56±17/2 | 70.7±14.2/3 | 38±0/1 | 61.7±15.5/3 | 44±0/1 |
| unknown | 0/4 (0) | 0/1 (0) | 0/40 (0) | 0/10 (0) | 0/5 (0) | 0/41 (0) | 0/12 (0) | 0/7 (0) | 0/30 (0) | 0/16 (0) |
| *Aftermath* | | | | | | | | | | |
| ambulatory | 0/4 (0) | 0/1 (0) | 0/40 (0) | 0/10 (0) | 0/5 (0) | 0/41 (0) | 0/12 (0) | 0/7 (0) | 0/30 (0) | 0/16 (0) |

| | | | | | | | | | | |
|---|---|---|---|---|---|---|---|---|---|---|
| inpatient | 0/4 (0) | 0/1 (0) | 0/40 (0) | 0/10 (0) | 1/5 (20) | 0/41 (0) | 0/12 (0) | 1/7 (14.3) | 1/30 (3.3) | 0/16 (0) |
| mortality | 0/4 (0) | 0/1 (0) | 0/40 (0) | 0/10 (0) | 0/5 (0) | 0/41 (0) | 0/12 (0) | 0/7 (0) | 0/30 (0) | 0/16 (0) |
| other | 0/4 (0) | 0/1 (0) | 3/40 (7.5) | 0/10 (0) | 0/5 (0) | 1/41 (2.4) | 1/12 (8.3) | 0/7 (0) | 1/30 (3.3) | 0/16 (0) |

Threat level from 0 (minimum) to 10 (maximum). Vac., vaccination. Ambulatory and inpatient refer to treatment requirement.

Suppl. Table 15:Dermatological symptoms

| | local rash | rash all over the body | swelling of lymph nodes | painful lymph nodes | extensive reddening of the skin | other skin discoloration | wheals/ hives | blisters with fluid/pus | petechiae | hematoma | dry skin | skin eczema | open skin areas | itching | other skin complaints |
|---|---|---|---|---|---|---|---|---|---|---|---|---|---|---|---|
| After any vac. | 78/242 (32.2) | 13/242 (5.4) | 69/242 (28.5) | 20/242 (8.3) | 5/242 (2.1) | 1/242 (0.4) | 14/242 (5.8) | 7/242 (2.9) | 11/242 (4.5) | 3/242 (1.2) | 37/242 (15.3) | 21/242 (8.7) | 4/242 (1.7) | 27/242 (11.2) | 8/242 (3.3) |
| After 1st vac. | 46/242 (19) | 7/242 (2.9) | 29/242 (12) | 7/242 (2.9) | 3/242 (1.2) | 1/242 (0.4) | 3/242 (1.2) | 5/242 (2.1) | 8/242 (3.3) | 2/242 (0.8) | 20/242 (8.3) | 10/242 (4.1) | 3/242 (1.2) | 16/242 (6.6) | 6/242 (2.5) |
| After 2nd vac. | 37/242 (15.3) | 6/242 (2.5) | 46/242 (19) | 11/242 (4.5) | 2/242 (0.8) | 1/242 (0.4) | 11/242 (4.5) | 4/242 (1.7) | 4/242 (1.7) | 2/242 (0.8) | 25/242 (10.3) | 14/242 (5.8) | 2/242 (0.8) | 18/242 (7.4) | 3/242 (1.2) |
| Threat level (mean±SD/N) | 0.8±1.3/78 | 1.9±2/13 | 0.6±1/69 | 0.4±0.7/20 | 0.4±0.5/5 | 0±0/1 | 1.9±1.9/14 | 0.6±1.1/7 | 0.7±1/11 | 0.3±0.6/3 | 0.4±1/36 | 0.9±1.7/21 | 0±0/4 | 0.7±1.5/27 | 0.4±0.5/8 |
| *Dosage* | | | | | | | | | | | | | | | |
| 3µg | 11/45 (24.4) | 0/7 (0) | 4/37 (10.8) | 0/11 (0) | 1/3 (33.3) | 1/1 (100) | 1/11 (9.1) | 1/5 (20) | 1/6 (16.7) | 0/2 (0) | 3/22 (13.6) | 4/16 (25) | 1/3 (33.3) | 2/15 (13.3) | 0/5 (0) |
| 5µg | 21/45 (46.7) | 5/7 (71.4) | 8/37 (21.6) | 0/11 (0) | 2/3 (66.7) | 0/1 (0) | 4/11 (36.4) | 3/5 (60) | 2/6 (33.3) | 1/2 (50) | 5/22 (22.7) | 3/16 (18.8) | 1/3 (33.3) | 8/15 (53.3) | 4/5 (80) |
| 10µg | 13/45 (28.9) | 2/7 (28.6) | 25/37 (67.6) | 11/11 (100) | 0/3 (0) | 0/1 (0) | 6/11 (54.5) | 1/5 (20) | 3/6 (50) | 1/2 (50) | 14/22 (63.6) | 9/16 (56.3) | 1/3 (33.3) | 5/15 (33.3) | 1/5 (20) |
| mixed/unknown | 33/78 (42.3) | 6/13 (46.2) | 32/69 (46.4) | 9/20 (45) | 2/5 (40) | 0/1 (0) | 3/14 (21.4) | 2/7 (28.6) | 5/11 (45.5) | 1/3 (33.3) | 15/37 (40.5) | 5/21 (23.8) | 1/4 (25) | 12/27 (44.4) | 3/8 (37.5) |
| *Age* | | | | | | | | | | | | | | | |
| <12 months | 3/78 (3.8) | 0/13 (0) | 0/69 (0) | 0/20 (0) | 0/5 (0) | 0/1 (0) | 0/14 (0) | 1/7 (14.3) | 0/11 (0) | 0/3 (0) | 0/37 (0) | 1/21 (4.8) | 0/4 (0) | 0/27 (0) | 0/8 (0) |
| 12 to <24 months | 23/78 (29.5) | 3/13 (23.1) | 10/69 (14.5) | 0/20 (0) | 0/5 (0) | 1/1 (100) | 2/14 (14.3) | 2/7 (28.6) | 1/11 (9.1) | 1/3 (33.3) | 5/37 (13.5) | 4/21 (19) | 2/4 (50) | 7/27 (25.9) | 0/8 (0) |
| 24 to <60 months | 52/78 (66.7) | 10/13 (76.9) | 59/69 (85.5) | 20/20 (100) | 5/5 (100) | 0/1 (0) | 12/14 (85.7) | 4/7 (57.1) | 10/11 (90.9) | 2/3 (66.7) | 32/37 (86.5) | 16/21 (76.2) | 2/4 (50) | 20/27 (74.1) | 8/8 (100) |
| *Beginning & duration* | | | | | | | | | | | | | | | |
| beginning (daysafter vaccination, mean±SD/N) | 2.1±1.4/77 | 1.6±0.9/12 | 1.3±1/69 | 1.1±0.2/19 | 1.8±0.8/5 | 1±0/1 | 2.1±0.9/14 | 1.7±0.8/7 | 2.3±1.9/11 | 1.7±0.6/3 | 2.9±1.9/37 | 2.6±1.7/20 | 3±2.2/4 | 2.1±1.6/27 | 2.1±1.8/8 |
| duration (days, mean±SD/N) | 5.2±7.6/73 | 13.8±16.7/12 | 4.5±7.6/63 | 2.2±1.1/18 | 3.3±3/5 | 2±0/1 | 5.2±7.8/13 | 9±15.1/5 | 2.6±1.3/10 | 5±3.5/3 | 18.5±20.7/24 | 19.2±17.3/15 | 5.5±2.1/2 | 6.3±7.9/22 | 9.9±10.8/8 |
| > 90 days | 0/78 (0) | 0/13 (0) | 1/69 (1.4) | 0/20 (0) | 0/5 (0) | 0/1 (0) | 0/14 (0) | 0/7 (0) | 0/11 (0) | 0/3 (0) | 0/37 (0) | 0/21 (0) | 0/4 (0) | 0/27 (0) | 0/8 (0) |

| | | | | | | | | | | | | | | | |
|---|---|---|---|---|---|---|---|---|---|---|---|---|---|---|---|
| ongoing (days, mean±SD/N) | 47±37.3/3 | 70±0/1 | 96±0/2 | 0±0/0 | 0±0/0 | 0±0/0 | 62±0/1 | 78.5±33.2/2 | 5±0/1 | 0±0/0 | 86.8±97.1/11 | 166.7±173.5/3 | 40±0/1 | 166.7±175.5/3 | 0±0/0 |
| unknown | 0/78 (0) | 0/13 (0) | 0/69 (0) | 0/20 (0) | 0/5 (0) | 0/1 (0) | 0/14 (0) | 0/7 (0) | 0/11 (0) | 0/3 (0) | 1/37 (2.7) | 0/21 (0) | 0/4 (0) | 0/27 (0) | 0/8 (0) |
| *Aftermath* | | | | | | | | | | | | | | | |
| ambulatory | 5/78 (6.4) | 3/13 (23.1) | 0/69 (0) | 0/20 (0) | 0/5 (0) | 0/1 (0) | 4/14 (28.6) | 2/7 (28.6) | 1/11 (9.1) | 0/3 (0) | 1/37 (2.7) | 2/21 (9.5) | 0/4 (0) | 1/27 (3.7) | 0/8 (0) |
| inpatient | 0/78 (0) | 0/13 (0) | 0/69 (0) | 0/20 (0) | 0/5 (0) | 0/1 (0) | 0/14 (0) | 0/7 (0) | 0/11 (0) | 0/3 (0) | 0/37 (0) | 0/21 (0) | 0/4 (0) | 0/27 (0) | 0/8 (0) |
| mortality | 0/78 (0) | 0/13 (0) | 0/69 (0) | 0/20 (0) | 0/5 (0) | 0/1 (0) | 0/14 (0) | 0/7 (0) | 0/11 (0) | 0/3 (0) | 0/37 (0) | 0/21 (0) | 0/4 (0) | 0/27 (0) | 0/8 (0) |
| other | 1/78 (1.3) | 1/13 (7.7) | 0/69 (0) | 0/20 (0) | 0/5 (0) | 0/1 (0) | 0/14 (0) | 0/7 (0) | 0/11 (0) | 0/3 (0) | 2/37 (5.4) | 0/21 (0) | 0/4 (0) | 1/27 (3.7) | 1/8 (12.5) |

Threat level from 0 (minimum) to 10 (maximum). Vac., vaccination. Ambulatory and inpatient refer to treatment requirement.

Suppl. Table 16: Non-BNT162b2 vaccines since Jan. 15th 2022, n (%)

| Vaccination | All N=2945 |
|---|---|
| Influenza | 1355 (47.2) |
| Meningococcal | 763 (26.6) |
| Measles/mumps/rubella with/without chickenpox | 609 (21.2) |
| Tetanus/diphtheria/pertussis and/or polio-pediatric polio | 558 (19.4) |
| Hepatitis A/B | 248 (8.6) |
| Human papillomavirus | 6 (0.2) |
| Other | 560 (19.5) |

Suppl. Table 17: Counts of missing data, n/N(%)

| | n=7806 |
|---|---|
| Female | 1 (0.0) |
| Male | 1 (0.0) |
| Diverse | 1 (0.0) |
| Age (years) (median, IQR) | 3 (0.0) |
| Height (cm) (median, IQR) | 108 (1.4) |
| Weight (kg) (median, IQR) | 65 (0.8) |
| Comorbidities (yes) | 0 (0) |
| Long-term Medication (yes) | 22 (0.3) |
| *Dosage* | |
| Dosage 1$^{st}$ vaccination | 566 (7.3) |
| Dosage 2$^{nd}$ vaccination | 492/7102 (6.9) |
| Dosage 3$^{rd}$ vaccination | 50/846 (5.9) |
| *Symptoms* | |
| Duration | 106 (1.4) |
| Local | 81 (1) |
| General | 102 (1.3) |
| Fever | 0 (0) |
| Musculoskeletal system | 120 (1.5) |
| Gastrointestinal | 120 (1.5) |
| Otolaryngological | 139 (1.8) |
| Pulmonary | 142 (1.8) |
| Cardiovascular | 139 (1.8) |
| Neurological | 157 (2) |
| Psychological | 157 (2) |
| Dermatological | 171 (2.2) |
| Number absent days | 333 (4.3) |
| Susceptibility to infections | 176 (2.3) |
| Threat rating scale | 266 (3.4) |
